# What can we learn from COVID-19 data by using epidemic models with unidentified infectious cases?

**DOI:** 10.1101/2021.06.16.21259019

**Authors:** Quentin Griette, Jacques Demongeot, Pierre Magal

**Affiliations:** Univ. Bordeaux, IMB, UMR 5251, F-33400 Talence, France; CNRS, IMB, UMR 5251, F-33400 Talence, France; Univ. Grenoble Alpes, AGEIS EA7407, F-38700 La Tronche, France

## Abstract

**Background:** The COVID-19 epidemic, which started in late December 2019 and rapidly spread throughout the world, was accompanied by an unprecedented release of reported case data. Our objective is to propose a fresh look at this data by coupling a phenomenological description to the epidemiological dynamics.

**Methods:** We use a phenomenological model to describe and regularize the data. This model can be matched by a single mathematical model reproducing the epidemiological dynamics with a time-dependent transmission rate. We provide a method to compute this transmission rate and reconstruct the changes in the social interactions between people as well as changes in host-pathogen interactions. This method is applied to the cumulative case data of 8 different geographic areas.

**Findings:** We reconstruct the transmission rate from the data, therefore we are in position to understand the contribution of the dynamical effects of social interactions (contacts between individuals) and the contribution of the dynamics of the epidemic. We deduce from the comparison of several instantaneous reproduction numbers that the social effects are the most important in the dynamic of COVID-19. We obtain an instantaneous reproduction number that stays below 3.5 from early beginning of the epidemic.

**Conclusion:** The instantaneous reproduction number staying below 3.5 implies that it is sufficient to vaccinate 71% of the population in each state or country considered in our study. Therefore assuming the vaccines will remain efficient against the new variants, and to be more confident it is sufficient to vaccinate 75 − 80% to get rid of COVID-19 in each state or country.

**Funding:** This research was funded by the Agence Nationale de la Recherche in France (Project name: MPCUII (PM) and (QG))

## Introduction

Since the first cases occurred in early December, 2019, the COVID-19 crisis has been accompanied by an unprecedented release of data. The first cluster of cases has been reported on December 31, 2019, by WHO^32^. Chinese authorities confirmed on January 7th that this cluster was caused by a novel coronavirus^31^. The disease then rapidly spread throughout the world; a case was identified in the U.S.A. as early as January 19, 2020, for instance^14^. According to the WHO database^1^, the first cases in Japan date back to January 14, in Italy to January 29 (even though the cluster of cases was announced on January 21, 2020^3^), in France to January 24, 2020, etc. The spread of the epidemic across countries was monitored and the data made publicly available at the international level by recognized scientific and political institutions such as WHO^1^ and the Johns Hopkins University^11^, who aggregated data released by national health agencies. To the extent of our knowledge, it is the first time in history that such detailed epidemiological data are made publicly available at a global scale; this opens new questions and challenges for the scientific community.

Modeling efforts in order to analyze and predict the dynamics of the epidemics were initiated from the start^21,16,30^. Forecasting the propagation of the epidemic is, in particular, a key challenge in infectious disease epidemiology. It has quickly become clear to medical doctors and epidemiologists that covert cases (asymptomatic or unreported infectious cases) play an important role in the spread of the COVID-19. An early description of an asymptomatic transmission in Germany was reported by Rothe et al.^28^. It was also observed on the Diamond Princess cruise ship in Yokohama in Japan by Mizumoto et al.^24^ that many of the passengers were tested positive to the virus, but never presented any symptoms. On the French aircraft carrier Charles de Gaulle, clinical and biological data for all 1739 crew members were collected on arrival at the Toulon harbor and during quarantine: 1121 crew members (64%) were tested positive for COVID-19 using RT-PCR, and among these, 24% were asymptomatic (see^6^ for more information). We also refer to Qiu^27^ for more information about this problem. Models accounting for asymptomatic transmission have been used in agreement with reported case data from the start of the epidemic^21,19,17,18^. The implementation of such models depends, however, on the *a priori* knowledge of some characteristic parameters of the host-pathogen interaction, among which the ascertainment rate. Nishiura and collaborators^25^ estimated this ascertainment rate to 9.2% on a 7.5-days detection window, based on testing data of repatriated Japanese nationals from Wuhan. This was corrected later to 44% for non-severe cases^26^. An early review of SARS-CoV-2 facts can be found in the work of Bar-On et al^5^.

In order to describe mathematically the spread of COVID-19, Liu et al.^21^ first took into account the contamination of susceptible individuals by contacts with unreported infectious individuals. In the same work a new method to use the number of reported cases in SIR models was also proposed. These method and model were extended in several directions by the same group^19,17,18,20^ to include non-constant transmission rates and a period of exposure. More recently the method was extended and successfully applied to a Japanese age-structured dataset by Griette, Magal and Seydi^13^. The method was also extended to investigate the predictability of the outbreak in several countries including China, South Korea, Italy, France, Germany and the United Kingdom by Liu, Magal and Webb^20^.

Another key parameter to understand the dynamics of the COVID-19 epidemic is the transmission rate, defined as the fraction of all possible contacts between susceptible and infected that effectively result in a new infection per unit of time. Estimating the average transmission rate is one of the most crucial challenges in the epidemiology of communicable diseases. In practice, many factors can influence the actual transmission rate, (i) the coefficient of susceptibility; (ii) the coefficient of virulence; (iii) the number of contacts per unit of time^23^; (iv) the environmental conductivity^9^.

Epidemic models with time dependent transmission rate have been considered in several article in the literature. The standard approach is to fix a function of time which depends on some parameters and to fix those parameters by using a best fit to the data. In Chowell et al.^7^ a specific form was chosen for the rate of transmission and applied to the Ebola outbreak in Congo. Huo et al^15^ used a predefined transmission rate which is depending on parameters. Here we are going the other way around. We reconstruct the transmission rate from the data by using the model without choosing a predefined function for the transmission rate. Such an approach have been used in the early 70s by London and Yorke^22,33^ who used a discrete time approximation model, and discussed the time dependent rate of transmission in the context of measles, chickenpox and mumps. More recently, several authors^4,10,12^ used both an explicit formula and algorithms to reconstruct the transmission rate. These studies allow us to understand that the regularization of the data is a difficult problem, and is crucial to reconstruct a meaningful time dependent transmission rate.

In this paper, we present a new method to compute the transmission rate from cumulative reported case data. While the use of a predefined transmission rate *τ* (*t*) as a function of time can lead to very nice fits of the data, here we are looking for a more intrinsic relationship between the data and the transmission rate. Therefore we propose a different approach and use a two-step procedure. Firstly, we use a *phenomenological model* to describe the data and extract the general trend of the epidemic dynamics while removing the insignificant noise. Secondly, we derive an explicit relationship between the phenomenological model and the transmission rate. In other words, we compute the transmission rate directly from the data. As a result we can reconstruct an estimation of the state of the population at each time, including covert cases. Our method also provides new indicators for the epidemiological dynamics that are related to the reproductive number.

## Methods

### Data sources

We used reported case data for 8 different geographic areas, namely the state of California, France, India, Israel, Japan, Peru, Spain, UK. Apart from the California state for which we used data from the COVID tracking project^2^, the reported case data were taken from the WHO database^1^.

### Data regularization

In order to reconstruct the time-dependent transmission rate, we regularized the time series of cumulative reported cases by fitting regular curves to the data. We first identified the epidemic waves for each of the 8 geographic areas. A Bernoulli-Verhulst curve was then fitted to each epidemic wave by using the Levenberg-Marquardt algorithm. We report the detailed output of the algorithm in the supplementary material, including confidence bounds on the parameters. The model was completed by filling the time windows between two waves with straight lines. Finally, in order to obtain a smooth model, we applied a Gaussian filter with a standard deviation of 7 days to the curve.

### Mathematical model

In order to reconstruct the transmission rate, we used the underlying mathematical model. A flowchart of this model is presented in Figure 1. The model itself includes 5 parameters whose values were taken from the literature: the average length of the noninfectious incubation period (1 day, (E)xposed); the average length of the infectious incubation period (3 days, (I)nfectious); the average length of the symptomatic period (7 days, (R)eported or (U)nreported); the ascertainment rate (0.8). An additional parameters appears in the initial condition and could not be computed from the data: the initial number of unreported individuals. The transmission rate was computed from the regularized data and the assumed parameters according to a methodology adapted from Demongeot et al^10^, which we describe in the supplementary material. The same mathematical model was used to compute the instantaneous reproduction number *R*_*e*_(*t*) and the quasi-instantaneous reproduction number 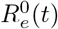. Another method (referred to as the “standard method”) was used to compute the instantaneous reproduction number from Cori et al^8^ 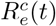. It corresponds to the green curves in the bottom row of Figures 4-5. This method is often described as purely statistical, even though it silently assumes an underlying time-independent evolution model on shifting time intervals.

**Figure 1:**
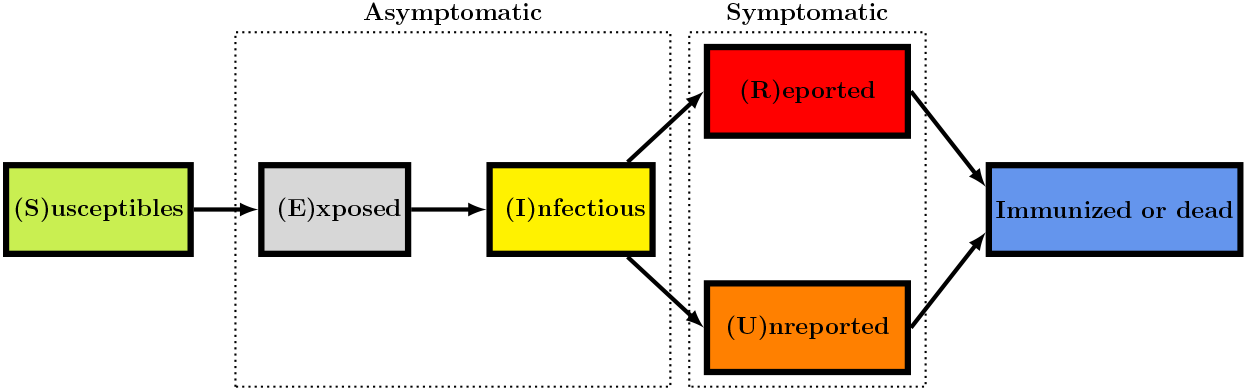
Flow chart for the model.

## Results

### Phenomenological model apply to COVID-19 data

Our method to regularize the data was applied to the 8 geographic areas. The resulting curves are presented in figures 2–3. The blue background color regions correspond to epidemic phases, and the yellow the background color regions to endemic phases. We added a plot of the daily number of cases (black dots) and the derivative of the regularized model for comparison, even though the daily number of cases is not used in the fitting procedure. The figures show in general an extremely good agreement between the time series of reported cases (top row, black dots) and the regularized model (top row, blue curve). The match between the daily number of cases (bottom row, black dots) and the derivative of the regularized model (bottom row, blue curve) is also very good, even though it is not a part of the optimization process. The relative error between the regularized curve and the data may be relatively high at the beginning of the epidemic because of the stochastic nature of the infection process and the small number of infected individuals, but quickly drops below 1% (see the supplementary material for more details).

**Figure 2:**
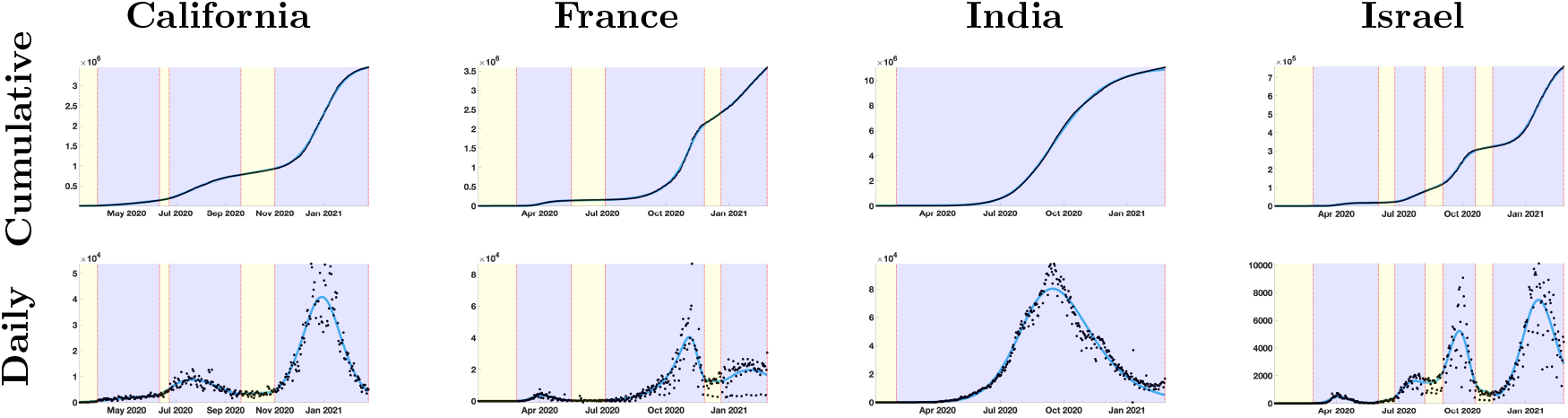
In the top four figures, we plot the cumulative number of reported cases (black dots) and the best fit of the phenomenological model (blue curve). In the bottom four figures, we plot the daily number of reported cases (black dots) and the first derivative of the phenomenological model (blue curve).

**Figure 3:**
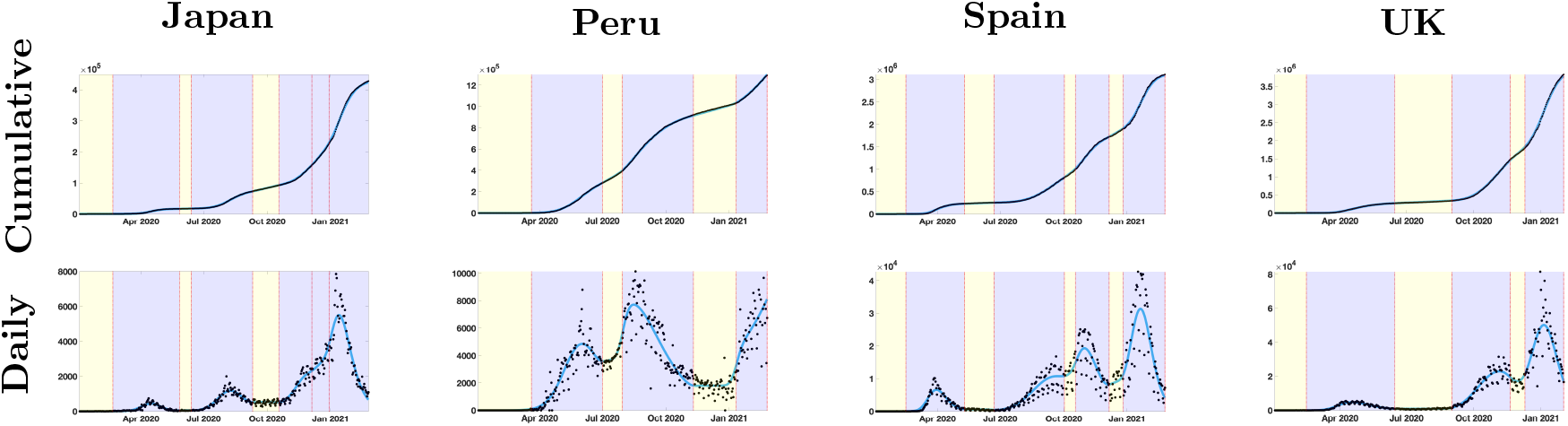
In the top four figures, we plot the cumulative number of reported cases (black dots) and the best fit of the phenomenological model (blue curve). In the bottom four figures, we plot the daily number of reported cases (black dots) and the first derivative of the phenomenological model (blue curve).

Non-identifiability of the ascertainment rate

Our analysis shows that it is hopeless to estimate the ascertainment rate by using time series of cumulative cases only. Indeed, a time-dependent transmission can be computed for any reasonable value of this rate, and fits the data perfectly. The same is true for other parameters of the mathematical model: the average duration of the non-infectious incubation period, the average duration of the infectious incubation period, the average duration of the symptomatic infectious period. Therefore, these parameters have to be estimated by statistical methods on samples of a different nature, like for the Diamond Princess cruise ship and the French aircraft carrier Charles de Gaulle.

### Bounds for the value of non-identifiable parameters

Even if some parameters of the mathematical model are not identifiable, we were able to gain some information on possible values for those parameters. Indeed, a mathematical model with a negative transmission rate *τ* (*t*) cannot be consistent with the real phenomenon. Therefore, parameter values which produce such negative values cannot be consistent with the data. Using this argument, we found that the average incubation period cannot exceed 8 days. The actual value of the upper bound is extremely variable across countries and epidemic waves. We report the values of the upper bound in Section 11 of the supplementary material.

### Instantaneous reproduction number computed for COVID-19 data

Our analysis allows us to compute the instantaneous transmission rate *τ* (*t*). We use this transmission rate to compute two different indicators of the epidemiological dynamics for each geographic areas, the instantaneous reproduction number and the quasi-instantaneous reproduction number. Both coincide with the basic reproduction number *R*_0_ at the first day of the epidemic. The instantaneous reproduction number at time *t, R*_*e*_(*t*), is the basic reproduction number corresponding to an epidemic starting at time *t* with a constant transmission rate equal to *τ* (*t*) and with an initial population of susceptibles composed of *S*(*t*) individuals (the number of susceptible individuals remaining in the population). The quasi-instantaneous reproduction number at time *t*, 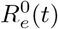, is the basic reproduction number corresponding to an epidemic starting at time *t* with a constant transmission rate equal to *τ* (*t*) and with an initial population of susceptibles composed of *S*_0_ individuals (the number of susceptible individuals at the start of the epidemic). The two indicators are represented for each geographic area in the top row of Figures 4 and 5 (black curve: instantaneous reproduction number; green curve: quasi-instantaneous reproduction number).

**Figure 4:**
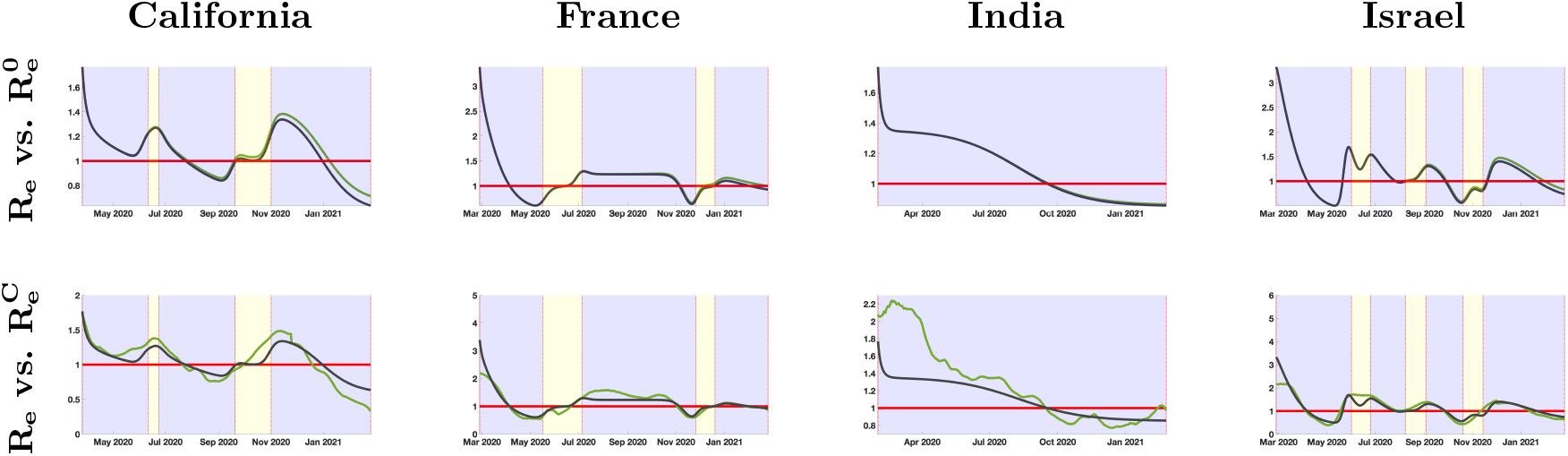
In the top four figures, we plot the instantaneous reproduction number R_e_(t) (in black) and the quasi instantaneous reproduction number 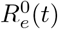 (in green). In the bottom four figures, we plot the instantaneous reproduction number R_e_(t) (in black) and the one obtained by the standard method^8,29^ 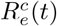 (in green).

**Figure 5:**
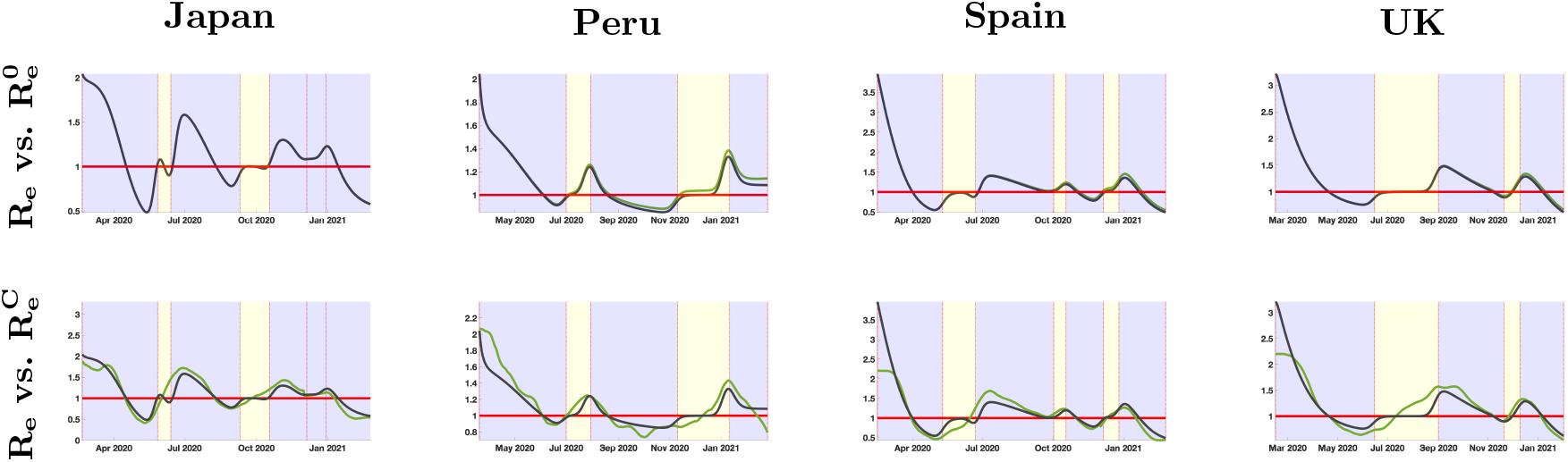
In the top four figures, we plot the instantaneous reproduction number R_e_(t) (in black) and the quasi instantaneous reproduction number 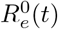 (in green). In the bottom four figures, we plot the instantaneous reproduction number R_e_(t) (in black) and the one obtained by the standard method^8,29^ 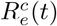 (in green).

Both indicators have a different interpretation. The instantaneous reproduction number indicates if, given the current state of the population, the epidemic tends to persist or die out in the long term (note that our model assumes that recovered individuals are perfectly immunized). The quasi-instantaneous reproduction number indicates if, the population being at its initial state, the epidemic tends to persist or die out in the long term. Also, it is directly proportional to the transmission rate and therefore allows to monitor its changes. Note that the value of 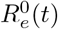 changed drastically between epidemic phases, revealing that *τ* (*t*) is far from constant. In any case, the difference between the two values starts to be visible in the figures one year after the start of the epidemic.

We also computed the reproduction number by using the method described in Cori et al^8^, which we denote 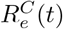. The precise implementation is described in the supplementary material. It is plotted in the bottom row of Figure 4 and Figure 5 (green curve), along with the instantaneous reproduction number *R*_*e*_(*t*) (green curve).

### Consequences for the vaccination

We observe that the value of the instantaneous and quasi-instantaneous reproduction numbers are both maximal at the beginning of the epidemic, when they coincide with the basic reproduction number. Based on this maximal value, our analysis provides a way to estimate the proportion of people that should be vaccinated in order to stop the propagation of the epidemic. Indeed, in most geographic areas in the present study, the instantaneous reproduction number is dominated by 3.5. Therefore if the proportion of vaccinated people is above *p* = 1 *—* 1*/*3.5 ≡ 0.71, the number of susceptible individuals would be below 1*/*3.5 × *S*_0_, and the instantaneous reproduction numbers would stay below 1.

## Discussion

In this article we presented a new phenomenological model to describe cumulative reported case data. This model allows us to handle multiple epidemic waves and fit very well the data for the eight geographic areas considered. The use of Bernoulli-Verhulst curves to fit an epidemic wave is not necessary. We expect that a number of different phenomenological models could be employed for the same purpose; however, our method has the advantage of involving a limited number of parameters. Moreover, the Bernoulli-Verhulst model leads to explicit algebraic formula for the compatibility conditions of non-identifiable parameters. It is far from obvious that the same computations can be carried out with other models. Our method also provides a very smooth curve with controlled upper-bound for the first (four) derivatives and we use the regularity obtained to compute the transmission rate. We refer to Demongeot, Griette and Magal^10^ for several examples of problems that may occur when using other methods to regularize the data (rolling weekly average, etc.)

The first goal of the article was to understand how to connect successive epidemic waves. As far as we know, this is new compared to the existing literature. A succession of epidemic waves separated by a short time period of random transmissions is regularly observed in the COVID-19 epidemic data. But several consecutive epidemic phases may happen without endemic transition. An illustration of this situation is provided by the case of Japan where the parameters of the Bernoulli-Verhulst model change three times during the last epidemic phases (without endemic interruption). Therefore we subdivide this last epidemic wave into three epidemic phases.

One other advantage of our method is the connection with an epidemiological model. Our study provides a way to explain the data by using a single epidemic model with a time-dependent transmission rate. More precisely, we find that there exists exactly one model that matches the best fit to the data. The fact that the transmission rate corresponding to the data is not constant is therefore meaningful. This means that the depletion of susceptible hosts due to natural epidemiological dynamics is not sufficient to explain the reduction in epidemic spread. Indeed, due to the social changes involving the distancing between individuals, the transmission rate should vary to take into account the changes in the number of contacts per unit of time. The variations in the observed dynamics of the number of cases mainly result from the modification of the people’s behavior. In other words, the social changes in the population have a stronger impact on the propagation of the disease than the pure epidemiological dynamics. By computing the transmission rate and the associated (effective) reproduction numbers we propose a new method to quantify those social changes. Other factors may also have an influence on the dynamics of the COVID-19 outbreak (temperature, humidity, etc.) and should be taken into account. However, the correlation between the dates of the waves and the mitigations measures imposed by local governments suggests that the former phenomenon takes a larger role in the epidemiological dynamics.

Precisely because it involves an epidemiological model, our method provides an alternative, robust way to compute indicators for the future behavior of the epidemic: the instantaneous and quasi-instantaneous reproductive numbers *R*_*e*_(*t*) and 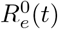. It is natural to compare them to an alternative in the literature, which is sometimes called “effective reproductive number”. The method of Cori et al^8^ is a popular framework to estimate its value. Compared to this standard method, however, our indicators perform better near the beginning of the epidemic and close to the last data point, and are less variable in time. That we require an *a priori* definition of epidemic waves can be considered as an advantage and a drawback. It is a drawback because the computed value of the indicator may slightly depend on the choice of the dates of the epidemic waves. On the other hand, this flexibility also allows to test different scenarios for the future evolution of the epidemic.

It appears from our results that the instantaneous reproduction number in almost every geographic areas considered is less than 3.5. Therefore, an efficient policy to get rid of the COVID-19 would be to vaccinate a fraction of 75 − 80% of the population. Once this threshold is reached, the situation should go back to normal in all the geographic areas considered in this study. This proportion can even be reduced at the expense of partially maintaining the social distancing and the other anti-COVID measures for a sufficiently long period of time.

Our method could also include several other features with few modifications. It is likely, for instance, that the vaccination of a large part of the population has an impact on the epidemiological dynamics, and this impact is not taken into account for the time being. Different distributions of serial intervals could be taken into account by replacing the mathematical model of ordinary differential equations by integral equations. What remains is that the coupling of a phenomenological model to describe the data, with an epidemiological model to take into account the nature of the underlying phenomenon, should provide us with a new, untapped source of information on the epidemic.

## Data Availability

no data were produced in the study

## Contributors

QG and PM conducted the analyses. QG and PM performed the literature search. QG did most of the numerical simulations. All authors contributed to the study design and drafting of the manuscript.

