## supplementary material for "What can we learn from COVID-19 data by using epidemic models with unidentified infectious cases?"

#### 1 Mathematical model

Many epidemiological models are based on the SIR or SEIR model which is classical in the context of epidemics. We refer to [23, 19] for the earliest articles devoted to such a question and to [1, 2, 6, 4, 5, 7, 9, 14, 16, 17, 20] for more models. In this article we will compare the following SEIUR model to the cumulative reported cases data

$$\begin{cases} S'(t) = -\tau(t)\left[I(t) + \kappa\, U(t)\right] S(t), \\ E'(t) = \tau(t)\left[I(t) + \kappa\, U(t)\right] S(t) - \alpha\, E(t), \\ I'(t) = \alpha\, E(t) - \nu\, I(t), \\ U'(t) = \nu\,(1-f)\, I(t) - \eta\, U(t), \\ R'(t) = \nu\, f\, I(t) - \eta\, R(t), \end{cases} \tag{1.1}$$

where at time $t$, $S(t)$ is the number of susceptible, $E(t)$ the number of exposed (not yet capable to transmit the pathogen), $I(t)$ the number of asymptomatic infectious, $R(t)$ the number of reported symptomatic infectious and $U(t)$ the number of unreported symptomatic infectious. This system is supplemented by initial data

$$S(t_0) = S_0,\ E(t_0) = E_0,\ I(t_0) = I_0,\ U(t_0) = U_0,\ \text{and } R(t_0) = R_0. \tag{1.2}$$

In this model, $\tau(t)$ is the rate of transmission, $1/\alpha$ is the average duration of the exposed period, $1/\nu$ is the average duration of the asymptomatic infectious period, and for simplicity we subdivide the class of symptomatic patients into the fraction $0 \leq f \leq 1$ of patients showing some severe symptoms, and the fraction $1-f$ of patients showing some mild symptoms assumed to be not detected. The quantity $1/\eta$ is the average duration of symptomatic infectious period.

The cumulative number of reported cases $\mathrm{CR}(t)$ is connected to the epidemic model by using the following relationship

$$\mathrm{CR}(t) = \mathrm{CR}_0 + \nu\, f\, \mathrm{CI}(t),\ \text{for } t \geq t_0, \tag{1.3}$$

where

$$\mathrm{CI}(t) = \int_{t_0}^{t} I(\sigma)\mathrm{d}\sigma. \tag{1.4}$$

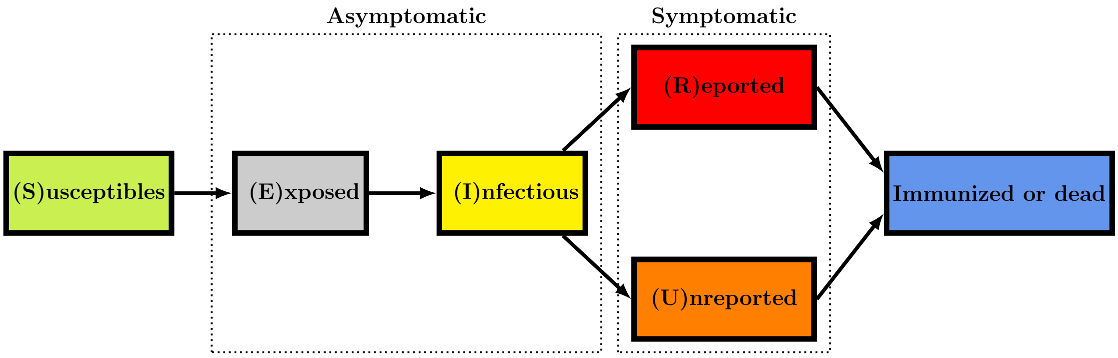


Figure 1: *Flow chart for the model.*

**Given and estimated parameters**

We assume that the following parameters of the model are known

$$S_0, U_0, R_0, f, \kappa, \alpha, \nu, \eta.$$

The goal of our method is to focus on the estimation of the three remaining parameters. Namely, knowing the above-mentioned parameters, we plan to identify

$$E_0, I_0, \tau(t).$$

**Computation of the rate of transmission**

The transmission rate is fully determined by the parameters $\kappa, \alpha, \nu, \eta, f, S_0, E_0, I_0, U_0$, and the data that are represented by the function $t \to \mathrm{CR}(t)$ and by using the three following equations

$$\tau(t) = \frac{1}{I(t) + \kappa U(t)} \times \frac{\mathrm{CE}''(t) + \alpha \mathrm{CE}'(t)}{E_0 + S_0 - \mathrm{CE}'(t) - \alpha \mathrm{CE}(t)}. \tag{1.5}$$

where

$$I(t) = \frac{\mathrm{CR}'(t)}{\nu\, f}, \tag{1.6}$$

$$\mathrm{CE}(t) = \frac{1}{\alpha\, \nu\, f} \left[ \mathrm{CR}'(t) - \nu\, f\, I_0 + \nu\, (\mathrm{CR}(t) - \mathrm{CR}_0) \right], \tag{1.7}$$

$$U(t) = e^{-\eta(t-t_0)} U_0 + \int_{t_0}^{t} e^{-\eta(t-s)} \frac{(1-f)}{f} \mathrm{CR}'(s) \mathrm{d}s. \tag{1.8}$$

#### Instantaneous reproduction number computed for COVID-19 data

In the standard SI epidemic model, we have only a single epidemic phase due the fact the epidemic exhausts the susceptible population. Here, the changes of regime (epidemic phase versus endemic phase) are partly due to the decay in the number of susceptible. But these changes are also influenced by the changes of the transmission rate. These changes of the transmission rate are due to the limitation of contacts between individuals or to changes in climate (in summer) or to other factors influencing transmissions.

In this section, we will observe that the main factor for the changes in the epidemic regimes are the changes in the transmission rate. In order to investigate this for the COVID-19 data, we use our method to compute the transmission rate and we consider the **instantaneous reproduction number**

$$R_e(t) = \frac{\tau(t)S(t)}{\eta\nu}(\eta + \nu(1-f)), \tag{1.9}$$

and the **quasi instantaneous reproduction number**

$$R_e^0(t) = \frac{\tau(t)S_0}{\eta\nu}(\eta + \nu(1-f)), \tag{1.10}$$

in which the transmission varies but the size of the susceptible population remains constant equal to $S_0$. We refer to the Appendix 10 for detailed computations to obtain the formula (1.9).

The comparison between $R_e(t)$ and $R_e^0(t)$, permits to understand the contribution of the decay of the susceptible population in the variations of $R_e(t)$. Another interesting aspect is that $R_e^0(t)$ is proportional to the transmission rate $\tau(t)$. Therefore plotting $R_e^0(t)$ permits to visualized the variation of $t \to \tau(t)$ only.

In the Results section, we computed three types of instantaneous reproduction numbers. The two first instantaneous reproduction numbers are obtained by using our method and combining the formula to compute the initial values $I_0$ in (2.18), $E_0$ in (2.20) as well as $\tau(t)$ in the formula (1.5). We observe that the changes between the first and the second notions of instantaneous reproduction numbers are small. This means that each epidemic phase is not stopping due to the fact that the susceptible individuals are exhausted by the epidemic. But the epidemic is stopping mainly due to the social changes. In the figure (c) the green curve is proportional to the transmission rate and we can see that the transmission rate is decaying due to social changes during epidemic phase and increase again when the new epidemic phase starts.

In the figure (d) for each country we compare the instantaneous reproduction numbers obtained by our method in black and the classical method of Cori et al [8] in green. We observe that the two methods are not the same at the beginning. That is because the method of Cori et al [8] is assuming that is not taking into account the initial values $I_0$ and $E_0$ while we do. Indeed the method Cori et al [8] is

### 2 Phenomenological model use for the multiple epidemic waves

In order represent the data we will use a phenomenological model to fit the curve of cumulative rate cases. Such an idea is not new, since it was already propose by Bernoulli [3] in 1760 in the context of smallpox epidemic. Here we use the so called Benoulli-Verhulst [21] model to describe the epidemic phase. Bernoulli [3] investigated an epidemic phase followed by an endemic phase. This appears clearly in the Figures 9 and 10 in [11]. Several works comparing cumulative reported case data and Bernoulli-Verhulst's model appear in the literature (see [15, 22, 24]). The Benoulli-Verhulst's is sometime called Richard's model while Richard's work came much more recently in 1959.

The phenomenological model deals with data series of new infectious cases decomposed into two types of successive phases, 1) endemic phases, followed by 2) epidemic ones.

**Endemic phase:** During the endemic phase, the dynamics of new cases appears to fluctuate around an average value independently of the number of cases. Therefore the average cumulative number of cases is given by

$$\text{CR}(t) = N_0 + (t - t_0) \times a, \text{ for } t \in [t_0, t_1], \tag{2.11}$$

where $t_0$ denotes the beginning of the endemic phase, and $a$ is the average value of daily number of new cases.

We assume that the average daily number of new cases is constant. Therefore the daily number of new cases is given by

$$\text{CR}'(t) = a. \tag{2.12}$$

**Epidemic phase:** In the epidemic phase, the new cases are contributing to produce second cases. Therefore the daily number of new cases is no longer constant, but varies with time as follows

$$\text{CR}(t) = N_{\text{base}} + \frac{e^{\chi(t-t_0)}N_0}{\left[1 + \frac{N_0^\theta}{N_\infty^\theta}\left(e^{\chi\theta(t-t_0)} - 1\right)\right]^{1/\theta}}, \text{ for } t \in [t_0, t_1]. \tag{2.13}$$

In other words, the daily number of new cases follows the Bernoulli-Verhulst [3, 21] equation. Namely, by setting

$$N(t) = \mathrm{CR}(t) - N_{\text{base}}, \tag{2.14}$$

we obtain

$$N'(t) = \chi\, N(t) \left[1 - \left(\frac{N(t)}{N_\infty}\right)^\theta\right], \tag{2.15}$$

completed with the initial value

$$N(t_0) = N_0.$$

In the model, $N_{\text{base}} + N_0$ corresponds to the value $\mathrm{CR}(t_0)$ of the cumulative number of cases at time $t = t_0$. The parameter $N_\infty + N_{\text{base}}$ is the maximal value of the cumulative reported cases after the time $t = t_0$. $\chi > 0$ is a Malthusian growth parameter, and $\theta$ regulates the speed at which the $\mathrm{CR}(t)$ increases to $N_\infty + N_{\text{base}}$.

**Regularized model:** Because the formula for $\tau(t)$ involves derivatives of the phenomenological model regularizing $\mathrm{CR}(t)$ (see equations (1.5)-(1.8)), we need to connect the phenomenological models of the different phases as smoothly as possible. Let us denote $t_0, \dots, t_n$ the $n+1$ breaking points of the model, that is to say, the times at which there is a transition between one phase and the next one. We let $\widetilde{\mathrm{CR}}(t)$ be the global model obtained by placing the phenomenological models for the different phases side by side. More precisely, $\widetilde{\mathrm{CR}}(t)$ is defined by (2.13) during an epidemic phase $[t_i, t_{i+1}]$, or during the initial phase $(-\infty, t_0]$ or the last phase $[t_n, +\infty)$. During an endemic phase, $\widetilde{\mathrm{CR}}(t)$ is defined by (2.11). The parameters are chosen so that the resulting global model $\widetilde{\mathrm{CR}}$ is continuous. We define the regularized model by using the convolution formula:

$$\mathrm{CR}(t) = \int_{-\infty}^{+\infty} \mathcal{G}(t-s) \times \widetilde{\mathrm{CR}}(s)\mathrm{d}s = (\mathcal{G} * \widetilde{\mathrm{CR}})(t), \tag{2.16}$$

where

$$\mathcal{G}(t) := \frac{1}{\sigma\sqrt{2\pi}} e^{-\frac{t^2}{2\sigma^2}},$$

is the Gaussian function with mean 0 and variance $\sigma^2$. The parameter $\sigma$ controls the trade-off between smoothness and precision: increasing $\sigma$ reduces the variations in $\mathrm{CR}(t)$ and reducing $\sigma$ reduces the distance between $\mathrm{CR}(t)$ and $\widetilde{\mathrm{CR}}(t)$. In any case the resulting function $\mathrm{CR}(t)$ is very smooth (as well as its derivatives) and close to the original model $\widetilde{\mathrm{CR}}(t)$ when $\sigma$ is not too large. In the result section, we fix $\sigma = 7$ days.

Numerically, we will need to compute some derivatives of $t \to \mathrm{CR}(t)$. Therefore it is convenient to take advantage of the convolution (2.16) and deduce that

$$\frac{d^n \mathrm{CR}(t)}{dt^n} = \int_{-\infty}^{+\infty} \frac{d^n \mathcal{G}(t-s)}{dt^n} \times \widetilde{\mathrm{CR}}(s)\mathrm{d}s, \tag{2.17}$$

for $n = 1, 2, 3$.

**Remark 1** *We tried several approaches to link an epidemic phase to the next endemic phase. So far this regularization procedure is the best one.*

#### 2.1 Computation of the initial value of the epidemic model

Based on (2.14), we can recover the initial number of asymptomatic infectious $I_0 = I(t_0)$ and the initial number of exposed $E_0 = E(t_0)$ for an epidemic phase starting at time $t_0$. Indeed by definition, we have $\mathrm{CR}'(t) = \nu f I(t)$ and therefore

$$I_0 = \frac{\mathrm{CR}'(t_0)}{\nu f} = \frac{\chi N_0 \left(1 - \left(\frac{N_0}{N_\infty}\right)^\theta\right)}{\nu f}.$$

**Estimated initial number of infected**

The initial number of asymptomatic infectious is given by

$$I_0 = \frac{\mathrm{CR}'(t_0)}{\nu\, f}. \tag{2.18}$$

In the special case of Bernoulli-Verhulst's model we obtain

$$I_0 = \frac{\chi}{\nu\, f}\, N_0 \left(1 - \left(\frac{N_0}{N_\infty}\right)^\theta\right). \tag{2.19}$$

By differentiating (2.15) we deduce that

$$\begin{aligned} N''(t) &= \chi\, N'(t) \left(1 - \left(\frac{N(t)}{N_\infty}\right)^\theta\right) - \frac{\chi\theta}{N_\infty^\theta}\, N(t)\, (N(t))^{\theta-1}\, N'(t) \\ &= \chi\, N'(t) \left(1 - \left(\frac{N(t)}{N_\infty}\right)^\theta\right) - \frac{\chi\theta}{N_\infty^\theta}\, (N(t))^{\theta}\, N'(t), \end{aligned}$$

therefore

$$\mathrm{CR}''(t) = N''(t) = \chi^2\, N(t) \left(1 - \left(\frac{N(t)}{N_\infty}\right)^\theta\right) \left(1 - (1+\theta) \left(\frac{N(t)}{N_\infty}\right)^\theta\right).$$

By using the third equation in (1.1) we obtain

$$E_0 = \frac{I'(t_0) + \nu I(t_0)}{\alpha} = \frac{\mathrm{CR}''(t_0) + \nu \mathrm{CR}'(t_0)}{\alpha} = \frac{N''(t_0) + \nu N'(t_0)}{\alpha}.$$

**Estimated initial number of exposed**

The initial number of exposed is given by

$$E_0 = \frac{\mathrm{CR}''(t_0) + \nu \mathrm{CR}'(t_0)}{\alpha}. \tag{2.20}$$

In the special case of Bernoulli-Verhulst's model we obtain

$$E_0 = \frac{\chi}{\alpha\, \nu\, f}\, N_0 \left(1 - \left(\frac{N_0}{N_\infty}\right)^\theta\right) \left(\chi + \nu - \chi\,(1+\theta) \left(\frac{N_0}{N_\infty}\right)^\theta\right). \tag{2.21}$$

### 3 Theoretical formula for $\tau(t)$

We first remark that the $S$-equation of model (1.1) can be written as

$$\frac{\mathrm{d}}{\mathrm{d}t} \ln(S(t)) = \frac{S'(t)}{S(t)} = -\tau(t)\big[I(t) + \kappa U(t)\big],$$

therefore by integrating between $t_0$ and $t$ we get

$$S(t) = S_0 \exp\left(-\int_{t_0}^t \tau(\sigma)\, \left[I(\sigma) + \kappa U(\sigma)\right] \mathrm{d}\sigma\right).$$

Next we plug the above formula for $S(t)$ into the $E$-equation of model (1.1) and obtain

$$\begin{aligned} E'(t) &= S_0 \exp\left(-\int_{t_0}^t \tau(\sigma)\, \left[I(\sigma) + \kappa U(\sigma)\right] \mathrm{d}\sigma\right) \tau(t)\, \left[I(t) + \kappa U(t)\right] - \alpha\, E(t) \\ &= -S_0 \frac{\mathrm{d}}{\mathrm{d}t} \left(-\int_{t_0}^t \tau(\sigma)\, \left[I(\sigma) + \kappa U(\sigma)\right] \mathrm{d}\sigma\right) \exp\left(-\int_{t_0}^t \tau(\sigma)\, \left[I(\sigma) + \kappa U(\sigma)\right] \mathrm{d}\sigma\right) - \alpha\, E(t), \end{aligned}$$

and by integrating this equation between $t_0$ and $t$ we obtain

$$E(t) = E_0 + S_0 \left[1 - \exp\left(-\int_{t_0}^{t} \tau(\sigma)\,\left[I(\sigma) + \kappa U(\sigma)\right] \mathrm{d}\sigma\right)\right] - \alpha \int_{t_0}^{t} E(\sigma)\mathrm{d}\sigma. \tag{3.22}$$

Define the cumulative numbers of exposed, infectious and unreported individuals by

$$\mathrm{CE}(t) := \int_{t_0}^{t} E(\sigma)\mathrm{d}\sigma,\ \mathrm{CI}(t) := \int_{t_0}^{t} I(\sigma)\mathrm{d}\sigma,\ \text{and}\ \mathrm{CU}(t) := \int_{t_0}^{t} U(\sigma)\mathrm{d}\sigma,$$

and note that $\mathrm{CE}'(t) = E(t)$. We can rewrite the equation (3.22) as

$$S_0 \exp\left(-\int_{t_0}^{t} \tau(\sigma)\,\left[I(\sigma) + \kappa U(\sigma)\right] \mathrm{d}\sigma\right) = E_0 + S_0 - \mathrm{CE}'(t) - \alpha\,\mathrm{CE}(t).$$

By taking the logarithm on both sides we obtain

$$\int_{t_0}^{t} \tau(\sigma)\,\left[I(\sigma) + \kappa U(\sigma)\right] \mathrm{d}\sigma = \ln(S_0) - \ln\left(E_0 + S_0 - \mathrm{CE}'(t) - \alpha\,\mathrm{CE}(t)\right),$$

and by differentiating with respect to $t$:

$$\tau(t) = \frac{1}{I(t) + \kappa U(t)} \times \frac{\mathrm{CE}''(t) + \alpha \mathrm{CE}'(t)}{E_0 + S_0 - \mathrm{CE}'(t) - \alpha\,\mathrm{CE}(t)}. \tag{3.23}$$

Therefore we have an explicit formula giving $\tau(t)$ as a function of $I(t)$, $U(t)$ and $\mathrm{CE}(t)$ and its derivatives. Next we explain how to identify those three remaining unknowns as a function of $\mathrm{CR}(t)$ and its derivatives. We first recall that, from (1.3), we have

$$\mathrm{CR}(t) = \mathrm{CR}(t_0) + \nu\, f\, \mathrm{CI}(t).$$

The $I$-equation of model (1.1) can be rewritten as

$$\alpha E(t) = I'(t) + \nu I(t),$$

and by integrating this equation between $t_0$ and $t$ we obtain

$$\alpha\,\mathrm{CE}(t) = \mathrm{CI}'(t) - I_0 + \nu\,\mathrm{CI}(t) = \frac{1}{\nu\, f}(\mathrm{CR}'(t) + \nu \mathrm{CR}(t) - \nu \mathrm{CR}(t_0)). \tag{3.24}$$

Finally by applying the variation of constants formula to the $U$-equation of system (1.1) we obtain

$$U(t) = e^{-\eta(t-t_0)}U_0 + \int_{t_0}^{t} e^{-\eta(t-s)}\nu\,(1-f)\,I(s)\mathrm{d}s = e^{-\eta(t-t_0)}U_0 + \int_{t_0}^{t} e^{-\eta(t-s)}\frac{1-f}{f}\mathrm{CR}'(s)\mathrm{d}s. \tag{3.25}$$

From these computations we deduce that $\tau(t)$ can be computed thanks to (3.23) from $\mathrm{CR}(t)$, $\alpha$, $\nu$, $\eta$, $\kappa$, $f$ and $U_0$. The following theorem is a precise statement of this result.

**Theorem 2** *Let $S_0 > 0$, $E_0 > 0$, $I_0 > 0$, $U_0 > 0$, $\mathrm{CR}_0 \geq 0$, $\alpha$, $\nu$, $\eta$ and $f > 0$ be given. Let $t \mapsto \tau(t) \geq 0$ be a given continuous function and $t \to I(t)$ be the second component of system (1.1). Let $\widehat{\mathrm{CR}} : [t_0, \infty) \to \mathbb{R}$ be a twice continuously differentiable function. Then*

$$\widehat{\mathrm{CR}}(t) = \mathrm{CR}_0 + \nu\, f \int_{t_0}^{t} I\,(s)\,ds, \forall t \geq t_0, \tag{3.26}$$

*if and only if $\widehat{\mathrm{CR}}$ satisfies*

$$\widehat{\mathrm{CR}}(t_0) = \mathrm{CR}_0, \tag{3.27}$$

$$\widehat{\mathrm{CR}}'(t_0) = \nu\, f\, I_0, \tag{3.28}$$

$$\widehat{\mathrm{CR}}''(t_0)+\nu\widehat{\mathrm{CR}}'(t_0)=\alpha\,\nu\,f\,E_0, \tag{3.29}$$

$$\widehat{\mathrm{CR}}'(t)>0,\forall t\geq t_0, \tag{3.30}$$

$$\nu f\,(E_0+S_0)-\left[\widehat{\mathrm{CR}}''(t)+\nu\widehat{\mathrm{CR}}'(t)\right]-\alpha\left[\widehat{\mathrm{CR}}'(t)-\nu\,f\,I_0+\nu\widehat{\mathrm{CR}}(t)\right]>0,\forall t\geq t_0, \tag{3.31}$$

*and $\tau(t)$ is given by*

$$\tau(t)=\frac{1}{\widehat{I}(t)+\kappa\widehat{U}(t)}\times\frac{\widehat{\mathrm{CE}}''(t)+\alpha\widehat{\mathrm{CE}}'(t)}{E_0+S_0-\widehat{\mathrm{CE}}'(t)-\alpha\widehat{\mathrm{CE}}(t)}, \tag{3.32}$$

*where*

$$\widehat{I}(t):=\frac{\widehat{\mathrm{CR}}'(t)}{\nu\,f}, \tag{3.33}$$

$$\widehat{\mathrm{CI}}(t):=\frac{1}{\nu\,f}\left[\widehat{\mathrm{CR}}(t)-\widehat{\mathrm{CR}}(t_0)\right], \tag{3.34}$$

$$\widehat{\mathrm{CE}}(t):=\frac{1}{\alpha}\left[\widehat{\mathrm{CI}}'(t)-I_0+\nu\,\widehat{\mathrm{CI}}(t)\right]=\frac{1}{\alpha\,\nu\,f}\left[\widehat{\mathrm{CR}}'(t)-\nu\,f\,I_0+\nu\left(\widehat{\mathrm{CR}}(t)-\mathrm{CR}_0\right)\right], \tag{3.35}$$

$$\widehat{U}(t):=e^{-\eta(t-t_0)}U_0+\int_{t_0}^{t}e^{-\eta(t-s)}\frac{(1-f)}{f}\widehat{\mathrm{CR}}'(s)\mathrm{d}s. \tag{3.36}$$

*Proof.* Assume first that $\widehat{\mathrm{CR}}(t)$ satisfies (3.26). Then by using the first equation of system (1.1) we deduce that

$$S_0\exp\left(-\int_{t_0}^{t}\tau(\sigma)\,\left[I(\sigma)+\kappa U(\sigma)\right]\mathrm{d}\sigma\right)=E_0+S_0-E(t)-\alpha\mathrm{CE}(t). \tag{3.37}$$

Therefore

$$\int_{t_0}^{t}\tau(\sigma)\,\left[I(\sigma)+\kappa U(\sigma)\right]\mathrm{d}\sigma=\ln\left[\frac{S_0}{E_0+S_0-E(t)-\alpha\mathrm{CE}(t)}\right]=\ln(S_0)-\ln\left[E_0+S_0-E(t)-\alpha\mathrm{CE}(t)\right],$$

and by taking the derivative on both side we obtain

$$\tau(t)\left[I(t)+\kappa U(t)\right]=\frac{E'(t)+\alpha E(t)}{E_0+S_0-E(t)-\alpha\mathrm{CE}(t)},$$

which is equivalent to

$$\tau(t)=\frac{E(t)}{I(t)+\kappa U(t)}\times\frac{\dfrac{E'(t)}{E(t)}+\alpha}{E_0+S_0-E(t)-\alpha\mathrm{CE}(t)}.$$

By using the fact that $E(t)=\mathrm{CE}'(t)$ and $I=\mathrm{CR}'(t)/(\nu f)$, we deduce (3.32). By differentiating (3.26), we get (3.28) and (3.30). (3.29) is a consequence of the $E$-component of equation (1.1). We get (3.31) by combining (3.37) and (3.35) (since $\widehat{\mathrm{CE}}(t)=\mathrm{CE}(t)$).

Conversely, assume that $\tau(t)$ is given by (3.31) and all the equations (3.27)–(3.36) hold. We define $\widehat{I}(t)=\widehat{\mathrm{CR}}'(t)/\nu f$ and $\widehat{\mathrm{CI}}(t)=\left(\widehat{\mathrm{CR}}(t)-\mathrm{CR}_0\right)/\nu f$. Then, by using (3.27), we deduce that

$$\widehat{\mathrm{CI}}(t)=\int_{t_0}^{t}\widehat{I}(\sigma)d\sigma, \tag{3.38}$$

and by using (3.28), we deduce

$$\widehat{I}(t_0) = I_0. \tag{3.39}$$

Moreover from (3.31) and $\widehat{I}(t) = \widehat{\text{CR}}'(t)/\nu f$, we deduce that

$$\tau(t) = \frac{1}{\widehat{I}(t) + \kappa \widehat{U}(t)} \times \frac{\widehat{\text{CE}}''(t) + \alpha \widehat{\text{CE}}'(t)}{E_0 + S_0 - \widehat{\text{CE}}'(t) - \alpha \widehat{\text{CE}}(t)}. \tag{3.40}$$

Multiplying (3.40) by $\widehat{I}(t) + \kappa \widehat{U}(t)$ and integrating, we obtain

$$\begin{aligned} \int_{t_0}^{t} \tau(\sigma) \left[\widehat{I}(\sigma) + \kappa \widehat{U}(\sigma)\right] \mathrm{d}\sigma = \; & \ln\left(E_0 + S_0 - \widehat{\text{CE}}'(t_0) - \alpha \widehat{\text{CE}}(t_0)\right) \\ & - \ln\left(E_0 + S_0 - \widehat{\text{CE}}'(t) - \alpha \widehat{\text{CE}}(t)\right), \end{aligned} \tag{3.41}$$

where the right-hand side is well-defined thanks to (3.31). By combining (3.27), (3.28) and (3.35) we obtain

$$\widehat{\text{CE}}(t_0) = 0, \tag{3.42}$$

and by taking the derivative in (3.35) we obtain

$$\widehat{\text{CE}}'(t_0) = \frac{1}{\alpha \, \nu \, f} \left[\widehat{\text{CR}}''(t) + \nu \widehat{\text{CR}}'(t)\right]$$

therefore by using (3.29) we deduce that

$$\widehat{\text{CE}}'(t_0) = E_0. \tag{3.43}$$

In particular $E_0 + S_0 - \widehat{\text{CE}}'(t_0) - \alpha \widehat{\text{CE}}(t_0) = S_0$ and, by taking the exponential of equation (3.41), we obtain

$$S_0 e^{-\int_{t_0}^{t} \tau(\sigma)\left[\widehat{I}(\sigma) + \kappa \widehat{U}(\sigma)\right] \mathrm{d}\sigma} = E_0 + S_0 - \widehat{\text{CE}}'(t) - \alpha \widehat{\text{CE}}(t),$$

which, differentiating both sides, yields

$$-S_0 e^{-\int_{t_0}^{t} \tau(\sigma)\left[\widehat{I}(\sigma) + \kappa \widehat{U}(\sigma)\right] \mathrm{d}\sigma} \tau(t) \left[\widehat{I}(t) + \kappa \widehat{U}(t)\right] = -\widehat{\text{CE}}''(t) - \alpha \widehat{\text{CE}}'(t) = -\widehat{E}'(t) - \alpha \widehat{E}(t),$$

and therefore

$$\widehat{E}'(t) = \tau(t) \widehat{S}(t) \left[\widehat{I}(t) + \kappa \widehat{U}(t)\right] - \alpha \widehat{E}(t), \tag{3.44}$$

where $\widehat{E}(t) := \widehat{\text{CE}}'(t)$ and $\widehat{S}(t) := S_0 e^{-\int_{t_0}^{t} \tau(\sigma)\left[\widehat{I}(\sigma) + \kappa \widehat{U}(\sigma)\right] \mathrm{d}\sigma}$. Differentiating the definition of $\widehat{S}(t)$, we get

$$\widehat{S}'(t) = -\left[\widehat{I}(t) + \kappa \widehat{U}(t)\right] \widehat{S}(t). \tag{3.45}$$

Next the derivative of (3.35) can be rewritten as

$$\widehat{I}'(t) = \frac{1}{\nu \, f} \widehat{\text{CR}}''(t) = \alpha \widehat{\text{CE}}'(t) - \nu \frac{1}{\nu \, f} \widehat{\text{CR}}'(t) = \alpha \widehat{E}(t) - \nu \widehat{I}(t). \tag{3.46}$$

Finally, differentiating (3.36) yields

$$\widehat{U}'(t) = \nu \, (1 - f) \, \widehat{I}(t) - \eta \widehat{U}(t). \tag{3.47}$$

By combining (3.44)–(3.47) we see that $\left(\widehat{S}(t), \widehat{E}(t), \widehat{I}(t), \widehat{U}(t)\right)$ satisfies (1.1) with the initial condition $\left(\widehat{S}(t_0), \widehat{E}(t_0), \widehat{I}(t_0), \widehat{U}(t_0)\right) = (S_0, E_0, I_0, U_0)$. By the uniqueness of the solutions of (1.1) for a given initial condition, we conclude that $\left(\widehat{S}(t), \widehat{E}(t), \widehat{I}(t), \widehat{U}(t)\right) = \left(S(t), E(t), I(t), U(t)\right)$. In particular $\text{CR}(t)$ satisfies (3.26). The proof is completed. ∎

**Remark 3** *The condition (3.31) is equivalent to say that*

$$E_0 + S_0 - \widehat{\text{CE}}'(t) - \alpha \widehat{\text{CE}}(t) > 0, \; \forall t \geq t_0.$$

**Remark 4** *The present computations originate for the work of Hadeler [13].*

### 4 Computing the explicit formula for $\tau(t)$ during an epidemic phase

In this section we assume that the curve of cumulative reported cases is given by the Bernoulli-Verhulst formula

$$N(t) := \mathrm{CR}(t) - N_{\mathrm{base}} = \frac{e^{\chi(t-t_0)} N_0}{\left[1 + \frac{N_0^\theta}{N_\infty^\theta}\left(e^{\chi\theta(t-t_0)} - 1\right)\right]^{1/\theta}}, \text{ for } t \in [t_0, t_1],$$

and we recall that

$$N'(t) = \chi N(t) \left(1 - \left(\frac{N(t)}{N_\infty}\right)^\theta\right).$$

Then we can compute an explicit formula for the components of the system (1.1). By definition we have

$$I(t) = \frac{\mathrm{CR}'(t)}{\nu f} = \frac{\chi}{\nu f} N(t) \left(1 - \left(\frac{N(t)}{N_\infty}\right)^\theta\right), \tag{4.48}$$

which gives

$$I'(t) = \frac{\mathrm{CR}''(t)}{\nu f} = \frac{\chi^2}{\nu f} N(t) \left(1 - \left(\frac{N(t)}{N_\infty}\right)^\theta\right) \left(1 - (1+\theta)\left(\frac{N(t)}{N_\infty}\right)^\theta\right),$$

so that by using the $I$-component in the system (1.1) we get

$$E(t) = \frac{1}{\alpha}\left(I'(t) + \nu I(t)\right) = \frac{1}{\alpha\nu f}\left(\mathrm{CR}''(t) + \nu \mathrm{CR}'(t)\right).$$

By integration, we get

$$\begin{aligned} \mathrm{CE}(t) &= \frac{1}{\alpha\,\nu\, f}\left[\left(\mathrm{CR}'(t) - \mathrm{CR}_0'\right) + \nu\left[\mathrm{CR}(t) - \mathrm{CR}(t_0)\right]\right], \\ &= \frac{1}{\alpha\,\nu\, f}\left[\chi N(t)\left(1 - \left(\tfrac{N(t)}{N_\infty}\right)^\theta\right) - \nu\, f\, I_0 + \nu\left[N(t) - N_0\right]\right], \\ &= \frac{1}{\alpha\,\nu\, f}\left[N(t)\left(\chi + \nu - \chi\left(\frac{N(t)}{N_\infty}\right)^\theta\right) - \nu\, f\, I_0 - \nu N_0\right]. \end{aligned}$$

and since

$$\nu\, f\, I_0 = \mathrm{CR}'(t_0) = N'(t_0) = \chi\, N_0 \left(1 - \left(\frac{N_0}{N_\infty}\right)^\theta\right),$$

hence we obtain

$$\mathrm{CE}(t) = \frac{1}{\alpha\,\nu\, f}\left[N(t)\left(\chi + \nu - \chi\left(\frac{N(t)}{N_\infty}\right)^\theta\right) - N_0\left(\chi + \nu - \chi\left(\frac{N_0}{N_\infty}\right)^\theta\right)\right].$$

Note also that we have explicit formulas for $E(t) = \mathrm{CE}'(t)$ and $E'(t) = \mathrm{CE}''(t)$,

$$E(t) = \mathrm{CE}'(t) = \frac{\chi}{\alpha\nu f}\left[N(t)\left(1 - \left(\frac{N(t)}{N_\infty}\right)^\theta\right)\left(\chi + \nu - \chi(1+\theta)\left(\frac{N(t)}{N_\infty}\right)^\theta\right)\right], \tag{4.49}$$

and

$$\begin{aligned} E'(t) = \mathrm{CE}''(t) = \frac{\chi^2}{\alpha\nu f} N(t)\left(1 - \left(\frac{N(t)}{N_\infty}\right)^\theta\right) \\ \times\left[\chi + \nu - (\chi(2+\theta) + \nu)(1+\theta)\left(\frac{N(t)}{N_\infty}\right)^\theta + \chi(1+\theta)(1+2\theta)\left(\frac{N(t)}{N_\infty}\right)^{2\theta}\right]. \end{aligned}$$

Next, recall the U-equation of (1.1), that is

$$U'(t) = \nu(1-f)I(t) - \eta U(t),$$

therefore by the variation of constant formula we have

$$\begin{aligned} U(t) &= e^{-\eta(t-t_0)}U(t_0) + \int_{t_0}^{t} e^{-\eta(t-s)}(1-f)\nu I(s)\mathrm{d}s \\ &= e^{-\eta(t-t_0}U_0 + \int_{t_0}^{t} e^{-\eta(t-s)}\frac{1-f}{f}CR'(s)\mathrm{d}s. \end{aligned} \tag{4.50}$$

**Explicit formula for the transmission rate during an epidemic phase**

The transmission rate $\tau(t)$ can be computed as

$$\tau(t) = \frac{\chi N(t)\left(1-\left(\frac{N(t)}{N_\infty}\right)^{\theta}\right)}{I(t)+\kappa U(t)} \times \frac{\left[A\left(\frac{N(t)}{N_\infty}\right)^{2\theta} - B\left(\frac{N(t)}{N_\infty}\right)^{\theta} + C\right]}{E_0+S_0-E(t)-\alpha \mathrm{CE}(t)}. \tag{4.51}$$

where

$$N(t) = \frac{e^{\chi(t-t_0)}N_0}{\left[1+\frac{N_0^\theta}{N_\infty^\theta}\left(e^{\chi\theta(t-t_0)}-1\right)\right]^{1/\theta}}, \text{ for } t \geq t_0, \tag{4.52}$$

and

$$A := \chi^2(1+\theta)(1+2\theta), \tag{4.53}$$
$$B := \chi(1+\theta)\big[\chi(2+\theta)+\nu+\alpha\big], \tag{4.54}$$
$$C := (\alpha+\chi)(\chi+\nu), \tag{4.55}$$

and $I(t)$ is given by (4.48), $E(t)$ by (4.49) and $U(t)$ by (4.50).

### 5 Compatibility conditions for the positivity of the transmission rate

Recall from (4.51):

$$\tau(t) = \frac{\chi N(t)\left(1-\left(\frac{N(t)}{N_\infty}\right)^{\theta}\right)}{I(t)+\kappa U(t)} \times \frac{\left[A\left(\frac{N(t)}{N_\infty}\right)^{2\theta} - B\left(\frac{N(t)}{N_\infty}\right)^{\theta} + C\right]}{E_0+S_0-E(t)-\alpha \mathrm{CE}(t)}.$$

Here we require that the numerator and the denominator of the last fraction stay positive for all times.

**Positivity of the numerator:** The model is compatible with the data if the transmission rate $\tau(t)$ stays positive for all times $t \in \mathbb{R}$. The numerator

$$p(N) := AN^2 - BN + C$$

is a second-order polynomial with $N \in (0,1)$. Let $\Delta := B^2 - 4AC$ the be the discriminant of $p(N)$. Since $p'(0) = -B < 0$ and

$$p'(N) = 0 \Leftrightarrow N = \frac{B}{2A}$$

we have two cases: 1) $\frac{B}{2A} \geq 1$; or 2) $0 < \frac{B}{2A} < 1$.

**Case 1:** If $\frac{B}{2A} \geq 1$, $p(N)$ is non-negative for all $N \in [0,1]$ if and only if

$$p(1) > 0 \Leftrightarrow A + C - B > 0. \tag{5.56}$$

Substituting $A$, $B$, $C$ by their expression, we get

$$\begin{aligned}A+C-B &= \chi^2(1+\theta)(1+2\theta)+(\alpha+\chi)(\chi+\nu)-\chi(1+\theta)(\chi(2+\theta)+\alpha+\nu)\\ &= \chi^2+2\chi^2\theta+\chi^2\theta+2\chi^2\theta^2+\alpha\chi+\alpha\nu+\chi^2+\chi\nu\\ &\quad -2\chi^2-\chi\theta-2\chi^2\theta-\chi^2\theta^2-\alpha\chi-\nu\chi-\alpha\chi\theta-\nu\chi\theta\\ &= \chi^2\theta^2+\alpha\nu-\alpha\chi\theta-\nu\chi\theta\\ &= (\alpha-\chi\theta)(\nu-\chi\theta).\end{aligned}$$

**Case 2:** If $\dfrac{B}{2A}<1$, $p(N)$ is non-negative for all $N\in[0,1]$ if and only if

$$p\left(\frac{B}{2A}\right)(1)>0 \Leftrightarrow \Delta<0 \Leftrightarrow B^2-4AC<0. \tag{5.57}$$

**Lemma 5** $\Delta<0 \Rightarrow A+C-B>0$.

*Proof.* We have

$$\Delta<0 \Rightarrow B^2-4AC\le(B-2A)^2 \Leftrightarrow B^2-4AC\le B^2-4AB+4A^2$$

and after simplifying the result follows. ■

**Positivity of the denominator:** Next we turn to the denominator in the expression of $\tau$, *i.e.* we want to ensure

$$E_0+S_0-E(t)-\alpha\mathrm{CE}(t)>0 \text{ for all } t\in\mathbb{R}. \tag{5.58}$$

We let $Y:=\dfrac{N(t)}{N_\infty}$ and remark that $E(t)+\alpha\mathrm{CE}(t)$ can be written as

$$\begin{aligned}E(t)+\alpha\mathrm{CE}(t) &= \frac{1}{\alpha\nu f}\big[\chi N_\infty Y(1-Y^\theta)(\chi+\nu-\chi(1+\theta)Y^\theta)\\ &\quad+\alpha N_\infty Y(\chi+\nu-\chi Y^\theta)-\alpha N_\infty Y_0(\chi+\nu-Y_0^\theta)\big]\\ &= \frac{N_\infty}{\alpha\nu f}Y\left[(\chi+\alpha)(\chi+\nu)-\chi(\alpha+\nu+\chi(2+\theta))Y^\theta+\chi^2(1+\theta)Y^{2\theta}\right]\\ &\quad-\frac{N_0}{\nu f}\left(\chi+\nu-Y_0^\theta\right),\end{aligned}$$

since we know that $A>0$. Therefore (5.58) becomes

$$Y\left[(\chi+\alpha)(\chi+\nu)-\chi(\chi+\nu+\chi(1+\theta)+\alpha)Y^\theta+\chi^2(1+\theta)Y^{2\theta}\right]\le\frac{\alpha\nu f}{N_\infty}\left[E_0+S_0+\frac{N_0}{\nu f}\left(\chi+\nu-\left(\frac{N_0}{N_\infty}\right)^\theta\right)\right].$$

We let

$$g(Y):=Y\left[(\chi+\alpha)(\chi+\nu)-\chi(\alpha+\nu+\chi(2+\theta))Y^\theta+\chi^2(1+\theta)Y^{2\theta}\right]$$

and notice that

$$g'(Y)=(\chi+\alpha)(\chi+\nu)-\chi(1+\theta)(\alpha+\nu+\chi(2+\theta))Y^\theta+\chi^2(1+2\theta)(1+\theta)Y^{2\theta},$$

is exactly $p(N):=AN^2-BN+C$.

Therefore assuming that $A+C-B>0$ the derivative $g'(Y)$ is positive and $g$ is strictly increasing. So we only have to check the final value $g(1)$. We get

$$\begin{aligned}\frac{\alpha\nu f}{N_\infty}\left(S_0+E_0+\frac{N_0}{\nu f}\left(\chi+\nu-\left(\frac{N_0}{N_\infty}\right)^\theta\right)\right) &\ge(\chi+\alpha)(\chi+\nu)-\chi(\alpha+\nu-\chi(2+\theta))+\chi^2(1+\theta)\\ &=\chi^2+\alpha\nu+\alpha\chi+\nu\chi+\chi^2+\chi^2\theta\\ &\quad-\alpha\chi-\nu\chi-2\chi^2-\chi^2\theta\\ &=\alpha\nu.\end{aligned}$$

**Compatibility for the positivity**

The SEIUR model is compatible with the data only when $\tau(t)$ stays positive for all $t \geq t_0$. Therefore the following two conditions should be met:

$$(\nu - \chi\theta)(\alpha - \chi\theta) \geq 0, \tag{5.59}$$

and

$$f + \frac{1}{\nu} \frac{N_0}{S_0 + E_0} \left( \chi + \nu - \left( \frac{N_0}{N_\infty} \right)^{\theta} \right) \geq \frac{N_\infty}{S_0 + E_0}. \tag{5.60}$$

### 6 Computing the explicit formula for $\tau(t)$ during an endemic phase

Recall that during an endemic phase the cumulative number of cases is assumed to be a line. Therefore,

$$\mathrm{CR}(t) = A\,(t - t_0) + B,$$

and

$$\mathrm{CR}'(t) = A \text{ and } \mathrm{CR}''(t) = 0.$$

Therefore

$$I(t) = \frac{\mathrm{CR}'(t)}{\nu f} = \frac{A}{\nu f}, \tag{6.61}$$

and

$$E(t) = \frac{I' + \nu I}{\alpha} = \frac{A}{\alpha f}. \tag{6.62}$$

Hence

$$\mathrm{CE}(t) = \frac{A}{\alpha f}\,(t - t_0)\,. \tag{6.63}$$

Moreover

$$U(t) = e^{-\eta(t-t_0)} U_0 + \int_{t_0}^{t} e^{-\eta(t-s)} \nu (1-f) I(s) ds,$$

and we obtain

$$U(t) = e^{-\eta(t-t_0)} U_0 + \frac{(1-f)A}{\eta f} \left( 1 - e^{-\eta(t-t_0)} \right). \tag{6.64}$$

By combining (1.5) and (6.61)-(6.64) we obtain the following explicit formula.

**Explicit formula for the transmission rate during an endemic phase**

The transmission rate $\tau(t)$ can be computed as

$$\tau(t) = \frac{1}{\frac{A}{\nu f} + \kappa \left( e^{-\eta(t-t_0)} U_0 + \frac{1-f}{\eta f} A \left( 1 - e^{-\eta(t-t_0)} \right) \right)} \times \frac{A}{f S_0 - A\,(t - t_0)}, \tag{6.65}$$

with the compatibility condition

$$t_0 \leq t < \frac{f S_0}{A} + t_0.$$

**Remark 6** *The above transmission rate corresponds to a constant number of daily infected $A$. Therefore it is not possible to maintain such a constant flux of new infected whenever the number of susceptible individuals is finite. The time $t = \frac{f S_0}{A} + t_0$ corresponds to the maximal time starting from $t_0$ during which we can maintain such a regime.*

### 7 Additional information for the results section

| Period | Interpretation | Parameters value | Method |
|---|---|---|---|
| $U_0$ | Number of unreported symptomatic infectious at time $t_0$ | 1 | Fixed |
| $R_0$ | Number of reported symptomatic infectious at time $t_0$ | 0 | Fixed |
| $\tau(t)$ | Transmission rate | (1.5)-(1.8) | Computed |
| $f$ | Fraction of reported symptomatic infectious | 0.8 | Fixed |
| $\kappa$ | Fraction of unreported symptomatic infectious capable to transmit the pathogen | 1 | Fixed |
| $1/\alpha$ | Average duration of the exposed period | 1 days | Fixed |
| $1/\nu$ | Average duration of the asymptomatic infectious period | 3 days | Fixed |
| $1/\eta$ | Average duration of the symptomatic infectious period | 7 days | Fixed |

Table 1: *In this table we list the values of the parameters of the epidemic model used for the simulations.*

#### 7.1 California

Figure 4 of the main text is devoted to the reproduction number of the model. The instantaneous reproduction number $t \to R_e(t)$ starts at around 1.8 on March 4 2020, then goes to a minimum above 1 on June 3 2020. Then we observe a peak during the endemic phase. During the second epidemic wave from June 23 to September 20 the $R_e(t)$ goes from 1.55 down to 0.81 on September 13 2020. Then we observe a new transmission period during the second endemic phase. During the last epidemic period $R_e(t)$ reaches a maximum at 1.39 on November 13 2020 then goes down to 0.75 on February 1 2021.

| Period | Interpretation | Parameters value | Method |
|---|---|---|---|
| $t_0$ | Time at which we started the epidemic model | Mar 26, 2020 | Fixed |
| $S_0$ | Number of susceptibles at time $t_0$ | $3.95 \times 10^7$ | Fixed |
| $E_0$ | Number of exposed at time $t_0$ | $7.91 \times 10^2$ | Computed |
| $I_0$ | Number of asymptomatic infectious at time $t_0$ | $2.06 \times 10^3$ | Computed |

Table 2: *In this table we list the values of the parameters of the epidemic model used for the simulations.*

**Compatibility condition between data and epidemic model**

By using the Californian data for the first, the second and the third epidemic waves, we get from (5.59) and (5.60) the following estimates for the average duration of the exposed and asymptomatic infectious periods and the fraction of reported cases

| | | |
|---|---|---|
| **First epidemic wave** | $\frac{1}{\alpha}$ and $\frac{1}{\nu} \leq \frac{1}{\chi\theta} = 5.23 \times 10^1$ days | $f \geq \frac{N_\infty}{S_0} = 8.21 \times 10^{-3}$ |
| **Second epidemic wave** | $\frac{1}{\alpha}$ and $\frac{1}{\nu} \leq \frac{1}{\chi\theta} = 2.52 \times 10^1$ days | $f \geq \frac{N_\infty}{S_0} = 2.08 \times 10^{-2}$ |
| **Third epidemic wave** | $\frac{1}{\alpha}$ and $\frac{1}{\nu} \leq \frac{1}{\chi\theta} = 1.54 \times 10^1$ days | $f \geq \frac{N_\infty}{S_0} = 6.72 \times 10^{-2}$ |

#### 7.2 France

Figure 4 of the main text is devoted to the reproduction number of the model. The instantaneous reproduction number $t \to R_e(t)$ starts at around 3.4 on February 27 2020 then goes to a minimum 0.6 on May 7 2020. The first confinement period started on March 17 and stopped on May 11 2020. Then we observe a transition during the first endemic phase. The second epidemic wave starts on July 5 2020 with a constant $R_e(t) = 1.23$ until October 15. $R_e(t) = 0.64$ goes to a minimum on November 21 2020. This corresponds to the second confinement period in France which goes from October 30 to December 15 2020. Then we observe a new endemic period during a transition to the third epidemic phase. During the last epidemic period $R_e(t)$ reaches a maximum at 1.1 on January 4 2021 then goes down to 0.92 on February 25 2021.

In Figure 4 of the main text the difference between $R_e(t)$ and $R_e^0(t)$ is almost not visible.

| Period | Interpretation | Parameters value | Method |
|---|---|---|---|
| $t_0$ | Time at which we started the epidemic model | Feb 27, 2020 | Fixed |
| $S_0$ | Number of susceptibles at time $t_0$ | $6.50 \times 10^7$ | Fixed |
| $E_0$ | Number of exposed at time $t_0$ | $4.27 \times 10^1$ | Computed |
| $I_0$ | Number of asymptomatic infectious at time $t_0$ | $6.30 \times 10^1$ | Computed |

Table 3: *In this table we list the values of the parameters of the epidemic model used for the simulations.*

**Compatibility condition between data and epidemic model**

By using the French data for the first, the second and the third epidemic waves, we get from (5.59) and (5.60) the following estimates for the average duration of the exposed and asymptomatic infectious periods and the fraction of reported cases

| | | |
|---|---|---|
| **First epidemic wave** | $\frac{1}{\alpha}$ and $\frac{1}{\nu} \leq \frac{1}{\chi\theta} = 1.17 \times 10^1$ days | $f \geq \frac{N_\infty}{S_0} = 2.19 \times 10^{-3}$ |
| **Second epidemic wave** | $\frac{1}{\alpha}$ and $\frac{1}{\nu} \leq \frac{1}{\chi\theta} = 4.15$ days | $f \geq \frac{N_\infty}{S_0} = 3.06 \times 10^{-2}$ |
| **Third epidemic wave** | $\frac{1}{\alpha}$ and $\frac{1}{\nu} \leq \frac{1}{\chi\theta} = 3.11 \times 10^1$ days | $f \geq \frac{N_\infty}{S_0} = 3.28 \times 10^{-2}$ |

#### 7.3 India

Figure 4 of the main text is devoted to the reproduction number of the model. The instantaneous reproduction number $t \to R_e(t)$ is decreasing from February 01, 2020 until February 25, 2021.

| Period | Interpretation | Parameters value | Method |
|---|---|---|---|
| $t_0$ | Time at which we started the epidemic model | Feb 01, 2020 | Fixed |
| $S_0$ | Number of susceptibles at time $t_0$ | $1.39 \times 10^9$ | Fixed |
| $E_0$ | Number of exposed at time $t_0$ | $4.29 \times 10^1$ | Computed |
| $I_0$ | Number of asymptomatic infectious at time $t_0$ | $1.12 \times 10^2$ | Computed |

Table 4: *In this table we list the values of the parameters of the epidemic model used for the simulations.*

**Compatibility condition between data and epidemic model**

By using the Indian data for the first single wave, we get from (5.59) and (5.60) the following estimates for the average duration of the exposed and asymptomatic infectious periods and the fraction of reported cases

| | | |
|---|---|---|
| **First epidemic wave** | $\frac{1}{\alpha}$ and $\frac{1}{\nu} \leq \frac{1}{\chi\theta} = 3.99 \times 10^1$ days | $f \geq \frac{N_\infty}{S_0} = 7.93 \times 10^{-3}$ |

#### 7.4 Israel

Figure 4 of the main text is devoted to the reproduction number of the model. The instantaneous reproduction number $t \to R_e(t)$ starts at around 3.3 on February 27, 2020 and then decreases to a global minimum at 0.5 on May 10, 2020. The Israeli government gradually imposed restrictions from the first cases, by limiting social gatherings on March 10, 2020. The lockdown measures were eased starting May 3, 2020 with the reopening of schools and ended on May 20 with the reopening of beaches and museums. The reproductive number $R_e(t)$ becomes greater than one on May 21 and stays above one until the period of July 2, 2020 to August 13 when it stays sligthly lower than one. The maximal value of $R_e(t)$ between May 21 and July 27 is 1.7. After August 13, there is a peak with maximum of 1.3 until September 29 2020 when the $R_e(t)$ becomes lower than one and stays below one until November 20 2020. The minimal value during this period is 0.6. There is a final peak of the value of $R_e(t)$ with maximal value at 1.4 on December 05 2020 after which the basic reproduction number decreases until February 25, 2021 to a value of 0.7. It becomes lower than one on January 21 2021.

| Period | Interpretation | Parameters value | Method |
|---|---|---|---|
| $t_0$ | Time at which we started the epidemic model | Feb 27, 2020 | Fixed |
| $S_0$ | Number of susceptibles at time $t_0$ | $8.74 \times 10^6$ | Fixed |
| $E_0$ | Number of exposed at time $t_0$ | 4.16 | Computed |
| $I_0$ | Number of asymptomatic infectious at time $t_0$ | 6.25 | Computed |

Table 5: *In this table we list the values of the parameters of the epidemic model used for the simulations.*

**Compatibility condition between data and epidemic model**

By using the Israeli data for the first, the second, the third and the fourth epidemic waves, we get from (5.59) and (5.60) the following estimates for the average duration of the exposed and asymptomatic infectious periods and the fraction of reported cases

| | | |
|---|---|---|
| **First epidemic wave** | $\frac{1}{\alpha}$ and $\frac{1}{\nu} \leq \frac{1}{\chi\theta} = 1.04 \times 10^1$ days | $f \geq \frac{N_\infty}{S_0} = 1.95 \times 10^{-3}$ |
| **Second epidemic wave** | $\frac{1}{\alpha}$ and $\frac{1}{\nu} \leq \frac{1}{\chi\theta} = 1.67 \times 10^1$ days | $f \geq \frac{N_\infty}{S_0} = 9.91 \times 10^{-3}$ |
| **Third epidemic wave** | $\frac{1}{\alpha}$ and $\frac{1}{\nu} \leq \frac{1}{\chi\theta} = 5.74$ days | $f \geq \frac{N_\infty}{S_0} = 2.69 \times 10^{-2}$ |
| **Fourth epidemic wave** | $\frac{1}{\alpha}$ and $\frac{1}{\nu} \leq \frac{1}{\chi\theta} = 1.71 \times 10^1$ days | $f \geq \frac{N_\infty}{S_0} = 5.57 \times 10^{-2}$ |

#### 7.5 Japan

Figure 4 of the main text is devoted to the reproduction number of the model. The instantaneous reproduction number $t \to R_e(t)$ decreases from 2.0 on January 20 2020 to 0.5 on May 13 2020. The basic reproductive number then increases again and becomes greater than one at the end of the first wave, on May 27 2020. There is an oscillation between the endemic phase May 27 2020 - June 13 2020, during which $R_e(t)$ takes values between 1.1 and 0.9. During the second epidemic phase (June 13, 2020 - September 13, 2020) the basic reproduction number first increases from the value 0.9 on June 13, 2020 to the maximal value of 1.6 on June 30, 2020, and then decreases back to a minimum of 0.8 on August 30. During the second endemic phase (September 10, 2020 - October 18, 2020) it stays close to the value 1.0. For the third and last epidemic phase, we identified three different regimes, with three different sets of parameters: October 18, 2020 - December 5, 2020, then December 05, 2020 - December 30, 2020, and finally December 30, 2020 - February 25, 2021. In the first period (October 18, 2020 - December 5, 2020) the basic reproduction number has a single peak at 1.3 on November 4, 2020 and the final value is 1.1 on December 05, 2020. During the second period (December 05, 2020 - December 30, 2020) the basic reproduction number increases until the value of 1.2 on December 20, 2020. In the last period, the basic reproduction number is decreasing, crosses the value 1.0 on January 14, 2021 and reaches the value of 0.6 on February 25, 2021.

| Period | Interpretation | Parameters value | Method |
|---|---|---|---|
| $t_0$ | Time at which we started the epidemic model | Feb 20, 2020 | Fixed |
| $S_0$ | Number of susceptibles at time $t_0$ | $1.26 \times 10^8$ | Fixed |
| $E_0$ | Number of exposed at time $t_0$ | 2.61 | Computed |
| $I_0$ | Number of asymptomatic infectious at time $t_0$ | 5.45 | Computed |

Table 6: *In this table we list the values of the parameters of the epidemic model used for the simulations.*

**Compatibility condition between data and epidemic model**

By using the Japanese data for the first, the second, the third, the fourth and the fifth epidemic waves, we get from (5.59) and (5.60) the following estimates for the average duration of the exposed and asymptomatic infectious periods and the fraction of reported cases

| | | |
|---|---|---|
| **First epidemic wave** | $\frac{1}{\alpha}$ and $\frac{1}{\nu} \leq \frac{1}{\chi\theta} = 8.18$ days | $f \geq \frac{N_\infty}{S_0} = 1.29 \times 10^{-4}$ |
| **Second epidemic wave** | $\frac{1}{\alpha}$ and $\frac{1}{\nu} \leq \frac{1}{\chi\theta} = 1.34 \times 10^{1}$ days | $f \geq \frac{N_\infty}{S_0} = 4.77 \times 10^{-4}$ |
| **Third epidemic wave** | $\frac{1}{\alpha}$ and $\frac{1}{\nu} \leq \frac{1}{\chi\theta} = 6.92$ days | $f \geq \frac{N_\infty}{S_0} = 7.22 \times 10^{-4}$ |
| **Fourth epidemic wave** | $\frac{1}{\alpha}$ and $\frac{1}{\nu} \leq \frac{1}{\chi\theta} = 7.17$ days | $f \geq \frac{N_\infty}{S_0} = 2.77 \times 10^{-3}$ |
| **Fifth epidemic wave** | $\frac{1}{\alpha}$ and $\frac{1}{\nu} \leq \frac{1}{\chi\theta} = 1.30 \times 10^{1}$ days | $f \geq \frac{N_\infty}{S_0} = 1.82 \times 10^{-3}$ |

#### 7.6 Peru

In Figure 2-(c) the difference between $R_e(t)$ and $R_e^0(t)$ is visible only starting from the second epidemic phase.

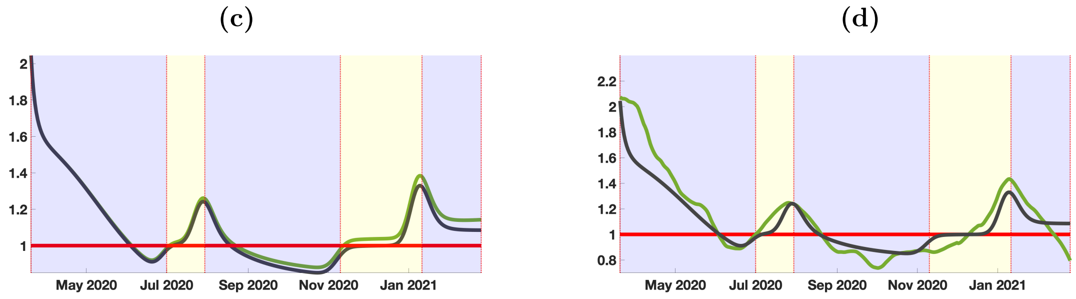


Figure 2: *In figure (c) we plot the instantaneous reproduction number $R_e(t)$ defined in (1.9) (in black) and the quasi instantaneous reproduction number $R_e^0(t)$ defined in (1.10) (in green) starting from the beginning of the first epidemic wave. In the figure (d), we plot the instantaneous reproduction number $R_e(t)$ defined in (1.9) (in black) and the one obtained by the standard method [8, 18] (in green). The transmission rate $\tau(t)$ is obtained by using the formula (1.5)-(1.8) together with the phenomenological model plotted in blue in Figure* **??***-(a). The parameters values used for the simulation are listed in Table 1 and Table 7.*

| Period | Interpretation | Parameters value | Method |
|---|---|---|---|
| $t_0$ | Time at which we started the epidemic model | Mar 20, 2020 | Fixed |
| $S_0$ | Number of susceptibles at time $t_0$ | $3.32 \times 10^7$ | Fixed |
| $E_0$ | Number of exposed at time $t_0$ | $1.64 \times 10^2$ | Computed |
| $I_0$ | Number of asymptomatic infectious at time $t_0$ | $3.85 \times 10^2$ | Computed |

Table 7: *In this table we list the values of the parameters of the epidemic model used for the simulations.*

**Compatibility condition between data and epidemic model**

By using the Peruvian data for the first, the second and the third epidemic waves, we get from (5.59) and (5.60) the following estimates for the average duration of the exposed and asymptomatic infectious periods and the fraction of reported cases

| | | |
|---|---|---|
| **First epidemic wave** | $\frac{1}{\alpha}$ and $\frac{1}{\nu} \leq \frac{1}{\chi\theta} = 2.20 \times 10^1$ days | $f \geq \frac{N_\infty}{S_0} = 1.09 \times 10^{-2}$ |
| **Second epidemic wave** | $\frac{1}{\alpha}$ and $\frac{1}{\nu} \leq \frac{1}{\chi\theta} = 3.47 \times 10^1$ days | $f \geq \frac{N_\infty}{S_0} = 2.32 \times 10^{-2}$ |
| **Third epidemic wave** | $\frac{1}{\alpha}$ and $\frac{1}{\nu} \leq \frac{1}{\chi\theta} = 2.01$ days | $f \geq \frac{N_\infty}{S_0} = 2.11 \times 10^{-1}$ |

#### 7.7 Spain

Figure 4 of the main text is devoted to the reproduction number of the model. The instantaneous reproduction number $t \to R_e(t)$ decreases from 3.9 on February 15, 2020 to 0.5 on April 29, 2020. The instantaneous reproduction number then increases again and reaches 0.7 at the end of the first wave, on May 10, 2020. During the first endemic period (May 10, 2020 - June 22, 2020) the instantaneous reproduction number increases to a plateau at 1.0 from May 27, 2020 to June 8, 2020 and then decreases to a valley at 0.9 on June 19, 2020. It does not significantly increase until June 22, 2020. During the second epidemic phase (June 22, 2020 - October 2nd, 2020) the instantaneous reproduction number first sharply increases from 0.9 on July 30, 2020 to the maximal value of 1.4 on July 11, 2020, and then slowly decreases to a valley at 1.0 on September 25, 2020. It does not significantly increase until October 2nd, 2020. During the second endemic phase (October 2nd, 2020 - October 18, 2021) it increases to reach a peak at 1.8 on October 18, 2020. During the third epidemic phase (October 18, 2020 - December 06, 2021), the instantaneous reproduction number decreases to a valley at 0.8 on November 23, 2020 and then increases to 1.0 on December 06, 2020. During the third endemic phase (December 06, 2020 - December 26, 2020), it increases to reach the value of 1.2. Finally during the fourth and last epidemic phase (December 26, 2020 - February 25, 2021), the instantaneous reproduction number reaches a peak at 1.4 on January 3rd, 2021 and then decreases to 0.5 on February 25.

| Period | Interpretation | Parameters value | Method |
|---|---|---|---|
| $t_0$ | Time at which we started the epidemic model | Feb 15, 2020 | Fixed |
| $S_0$ | Number of susceptibles at time $t_0$ | $3.95 \times 10^7$ | Fixed |
| $E_0$ | Number of exposed at time $t_0$ | 5.10 | Computed |
| $I_0$ | Number of asymptomatic infectious at time $t_0$ | 6.87 | Computed |

Table 8: *In this table we list the values of the parameters of the epidemic model used for the simulations.*

**Compatibility condition between data and epidemic model**

By using the Spanish data for the first, the second, the third and the fourth epidemic waves, we get from (5.59) and (5.60) the following estimates for the average duration of the exposed and asymptomatic infectious periods and the fraction of reported cases

| | | |
|---|---|---|
| **First epidemic wave** | $\frac{1}{\alpha}$ and $\frac{1}{\nu} \leq \frac{1}{\chi\theta} = 1.05 \times 10^1$ days | $f \geq \frac{N_\infty}{S_0} = 5.87 \times 10^{-3}$ |
| **Second epidemic wave** | $\frac{1}{\alpha}$ and $\frac{1}{\nu} \leq \frac{1}{\chi\theta} = 2.81 \times 10^1$ days | $f \geq \frac{N_\infty}{S_0} = 2.50 \times 10^{-2}$ |
| **Third epidemic wave** | $\frac{1}{\alpha}$ and $\frac{1}{\nu} \leq \frac{1}{\chi\theta} = 1.58 \times 10^1$ days | $f \geq \frac{N_\infty}{S_0} = 2.49 \times 10^{-2}$ |
| **Fourth epidemic wave** | $\frac{1}{\alpha}$ and $\frac{1}{\nu} \leq \frac{1}{\chi\theta} = 9.84$ days | $f \geq \frac{N_\infty}{S_0} = 3.29 \times 10^{-2}$ |

#### 7.8 United Kingdom

Figure 4 of the main text is devoted to the reproduction number of the model. The instantaneous reproduction number $t \to R_e(t)$ decreases from 3.2 on February 15, 2020 to a valley at 0.8 on June 1st,

2020. Then it increases to 0.9 at the end of the first epidemic phase on June 15, 2020. During the first endemic period (June 15, 2020 - September 1st, 2020) the instantaneous reproduction number increases to a plateau at 1.0 from June 21, 2020 to August 23, 2020 and then increases to 1.4 on September 1st, 2020. During the second epidemic phase (September 1st, 2020 - November 20, 2020) the instantaneous reproduction number first increases to a peak at 1.5 on September 9, 2020 and then decreases to 0.9 on November 20, 2020. During the second endemic phase (November 20, 2020 - December 10, 2021) it increases to 1.2. Finally during the third and last epidemic phase (December 10, 2020 - February 25, 2021), the instantaneous reproduction number reaches a peak at 1.3 on December 16, 2020 and then decreases to 0.6 on February 25.

| Period | Interpretation | Parameters value | Method |
|---|---|---|---|
| $t_0$ | Time at which we started the epidemic model | Feb 15, 2020 | Fixed |
| $S_0$ | Number of susceptibles at time $t_0$ | $6.81 \times 10^7$ | Fixed |
| $E_0$ | Number of exposed at time $t_0$ | 3.41 | Computed |
| $I_0$ | Number of asymptomatic infectious at time $t_0$ | 5.15 | Computed |

Table 9: *In this table we list the values of the parameters of the epidemic model used for the simulations.*

**Compatibility condition between data and epidemic model**

By using the data from Great Britain for the first, the second and the third epidemic waves, we get from (5.59) and (5.60) the following estimates for the average duration of the exposed and asymptomatic infectious periods and the fraction of reported cases

| | | |
|---|---|---|
| **First epidemic wave** | $\frac{1}{\alpha}$ and $\frac{1}{\nu} \leq \frac{1}{\chi\theta} = 2.06 \times 10^1$ days | $f \geq \frac{N_\infty}{S_0} = 4.20 \times 10^{-3}$ |
| **Second epidemic wave** | $\frac{1}{\alpha}$ and $\frac{1}{\nu} \leq \frac{1}{\chi\theta} = 3.14 \times 10^1$ days | $f \geq \frac{N_\infty}{S_0} = 3.15 \times 10^{-2}$ |
| **Third epidemic wave** | $\frac{1}{\alpha}$ and $\frac{1}{\nu} \leq \frac{1}{\chi\theta} = 1.08 \times 10^1$ days | $f \geq \frac{N_\infty}{S_0} = 3.56 \times 10^{-2}$ |

### 8 Plot of the multiple Bernoulli-Verhuslt's models fitted to each epidemic phase

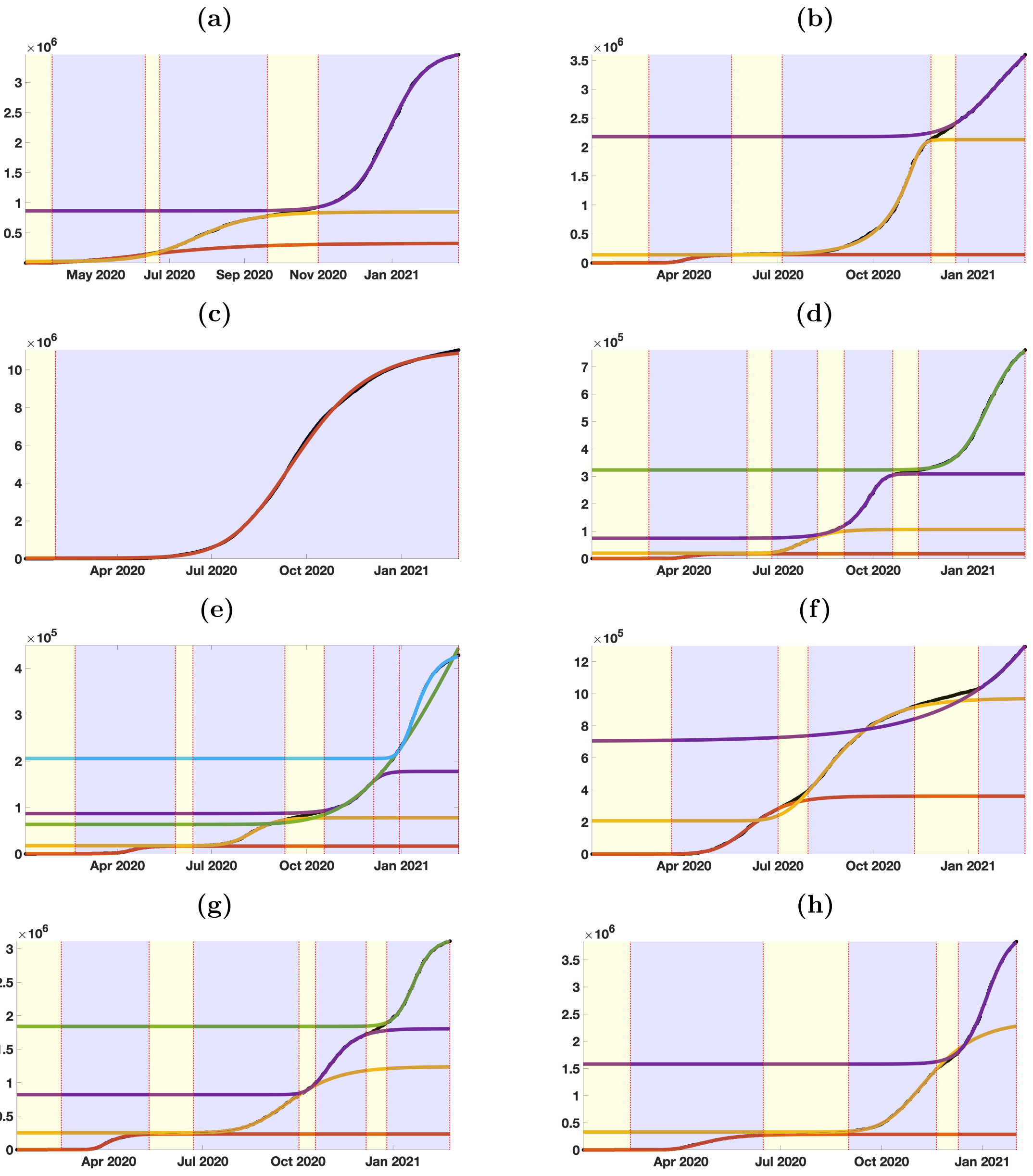


Figure 3: *In this figure, we plot the cumulative number of cases for California (black dots) and the best fit of Bernoulli-Verhulst for each epidemic wave for (a) California; (b) France; (c) India; (d) Israel; (e) Japan; (f) Peru; (g) Spain; (h) United Kingdom.*

### 9 Dependency with respect to the parameters for the French data

**Influence of $f$ on basic reproduction number:**

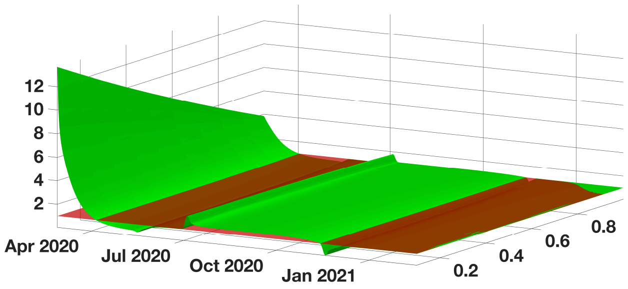


Figure 4: *In this figure we plot $(t, f) \to R_e(t)$ defined in (1.9) when $t$ varies from January 03 2020 to January 04 2021 and $f$ varies from* 0.1 *to* 1.

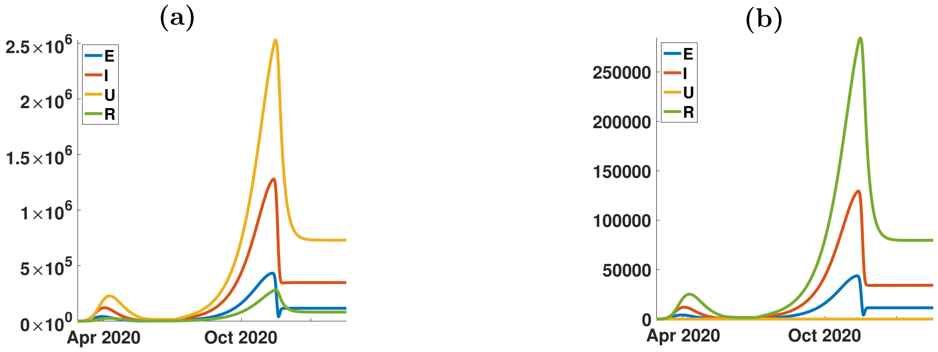


Figure 5: *In this figure we explore the influence of the parameter $f$ on the solution of model (1.1). The figure (a) corresponds to $f = 0.1$ and figure (b) corresponds to $f = 1$. The remaining parameters are unchanged.*

**Influence of $\kappa$ on basic reproduction number:**

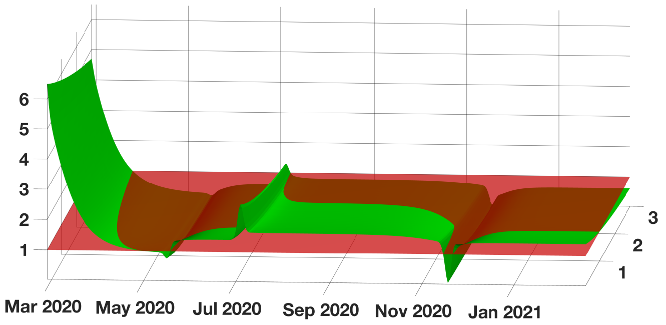


Figure 6: *In this figure we plot $(t, \kappa) \to R_e(t)$ defined in (1.9) when $t$ varies from January 03 2020 to January 04 2021 and $\kappa$ varies from* 0.1 *to* 3.

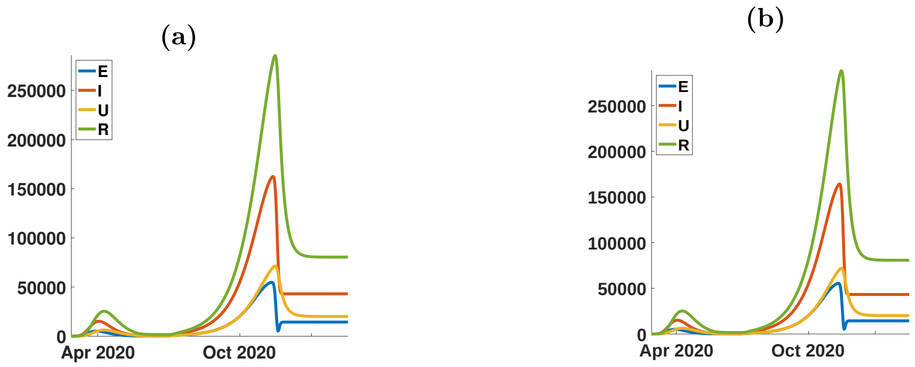


Figure 7: *In this figure we explore the influence of the parameter $f$ on the solution of model (1.1). The figure (a) corresponds to $\kappa = 0.1$ and figure (b) corresponds to $\kappa = 3$. The remaining parameters are unchanged.*

**Influence of $\nu$ on basic reproduction number:**

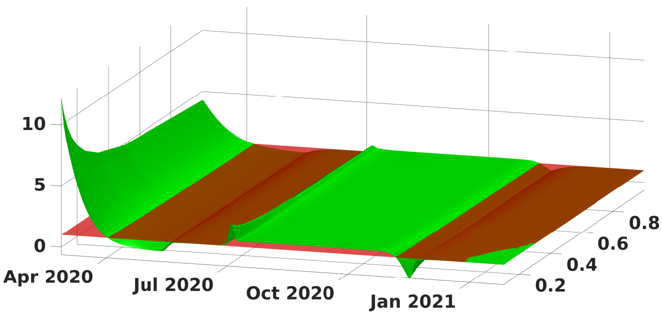


Figure 8: *In this figure we plot $(t, \nu) \to R_e(t)$ defined in (1.9) when $t$ varies from January 03 2020 to January 04 2021 and $\nu$ varies from 0.1 to 1 (or equivalently $1/\nu$ varies from 10 days to 1 day).*

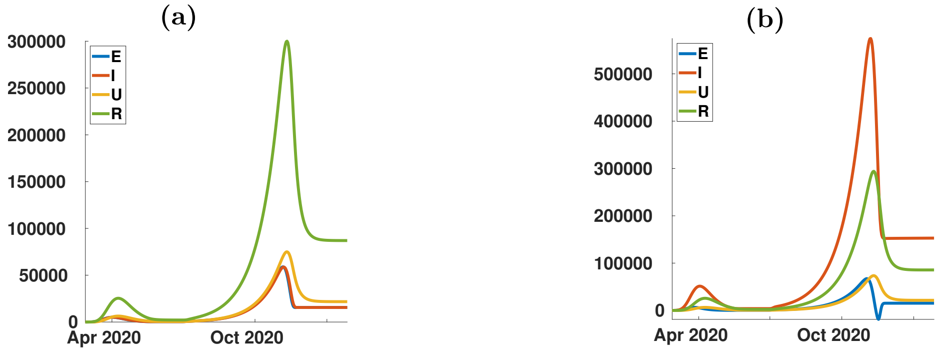


Figure 9: *In this figure we explore the influence of the parameter $1/\nu$ on the solution of model (1.1). The figure (a) corresponds to $1/\nu = 1$ and figure (b) corresponds to $1/\nu = 10$. The remaining parameters are unchanged.*

**Influence of $\eta$ on basic reproduction number:**

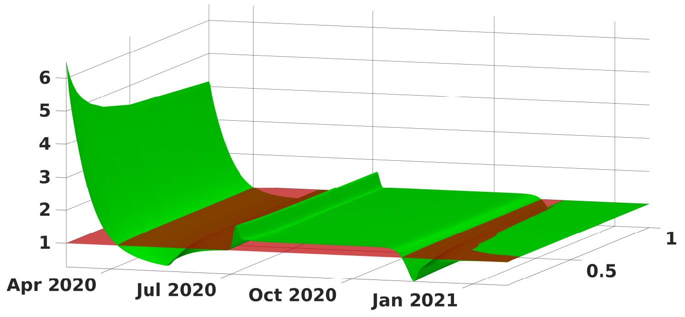


Figure 10: *In this figure we plot $(t, \eta) \to R_e(t)$ defined in (1.9) when $t$ varies from January 03 2020 to January 04 2021 and $\eta$ varies from* 0.1 *to* 1 *(or equivalently* $1/\eta$ *varies from* 10 *days to* 1 *day).*

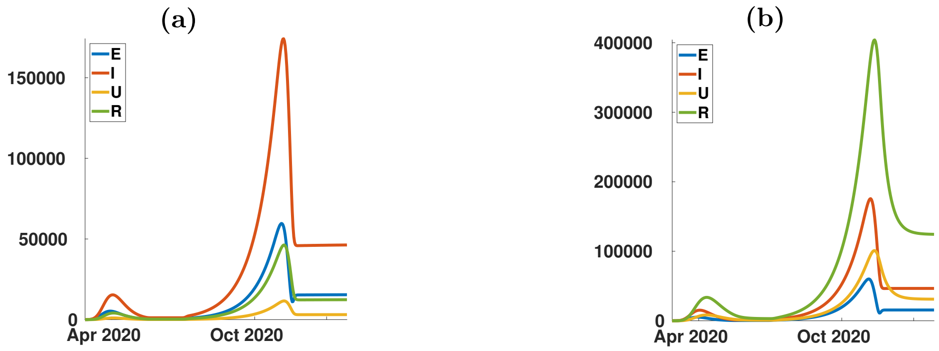


Figure 11: *In this figure we explore the influence of the parameter $f$ on the solution of model (1.1). The figure (a) corresponds to* $1/\eta = 1$ *days and figure (b) corresponds to* $1/\eta = 10$ *days. The remaining parameters are unchanged.*

**Influence of $\alpha$ on basic reproduction number:**

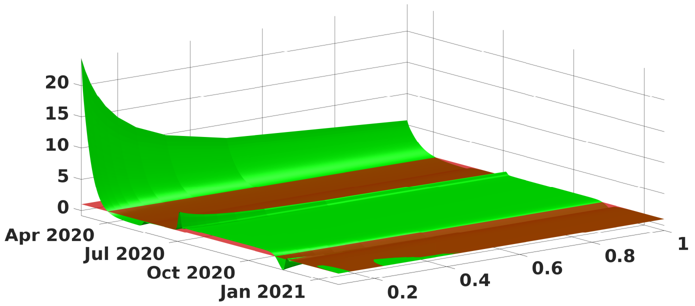


Figure 12: *In this figure we plot $(t, \alpha) \to R_e(t)$ defined in (1.9) when $t$ varies from January 03 2020 to January 04 2021 and $\alpha$ varies from* 0.1 *to* 1 *(or equivalently* $1/\alpha$ *varies from* 10 *days to* 1 *day).*

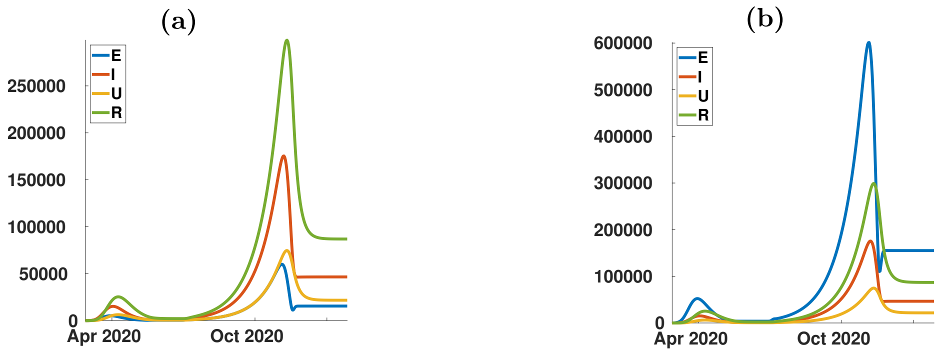


Figure 13: *In this figure we explore the influence of the parameter $f$ on the solution of model (1.1). The figure (a) corresponds to $1/\alpha = 1$ days and figure (b) corresponds to $1/\alpha = 10$ days. The remaining parameters are unchanged.*

### 10 Computing $R_0$

The basic reproduction number $R_0$ can be computed for the SEIUR model by the formula (see [10, 12])

$$R_0 = \rho(FV^{-1}),$$

where $F$ is the matrix containing new infections and $V$ contains the rates of transfer between classes:

$$F := \begin{pmatrix} 0 & \tau S & \tau\kappa S & 0 \\ 0 & 0 & 0 & 0 \\ 0 & 0 & 0 & 0 \\ 0 & 0 & 0 & 0 \end{pmatrix}, \qquad V := \begin{pmatrix} \alpha & 0 & 0 & 0 \\ -\alpha & \nu & 0 & 0 \\ 0 & -\nu(1-f) & \eta & 0 \\ 0 & \nu(1-f) & 0 & \eta \end{pmatrix},$$

see [10] and [12] for details. Therefore

$$V^{-1} = \begin{pmatrix} 1/\alpha & 0 & 0 & 0 \\ 1/\nu & 1/\nu & 0 & 0 \\ (1-f)/\eta & (1-f)/\eta & 1/\eta & 0 \\ f/\eta & f/\eta & 0 & 1/\eta \end{pmatrix}, FV^{-1} = \frac{\tau S}{\eta\nu} \begin{pmatrix} \eta + \kappa\nu(1-f) & \eta + \kappa\nu(1-f) & \kappa\nu & 0 \\ 0 & 0 & 0 & 0 \\ 0 & 0 & 0 & 0 \\ 0 & 0 & 0 & 0 \end{pmatrix}.$$

It follows that

$$R_0 = \frac{\tau S}{\eta\nu}\big(\eta + \kappa\nu(1-f)\big). \tag{10.66}$$

### 11 Relative error of the fitted curve compared to the data in each geographic area

#### 11.1 California

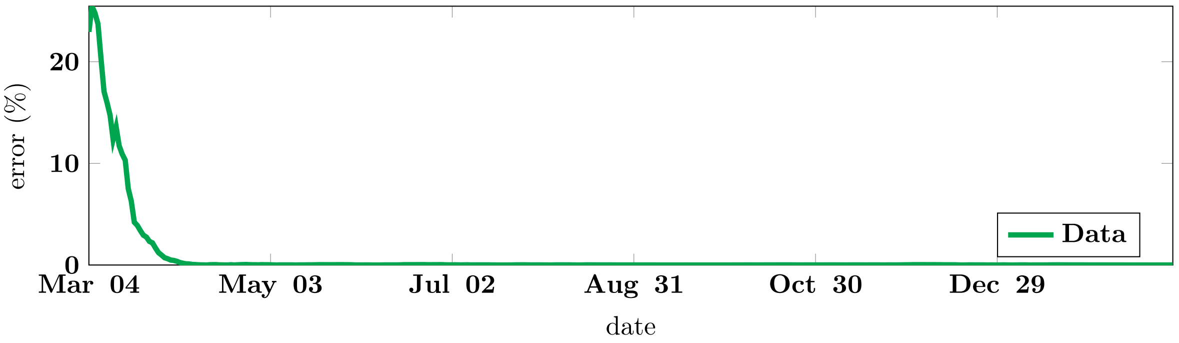


Figure 14: Relative error between the data and the model for California State, expressed in percent.

#### 11.2 France

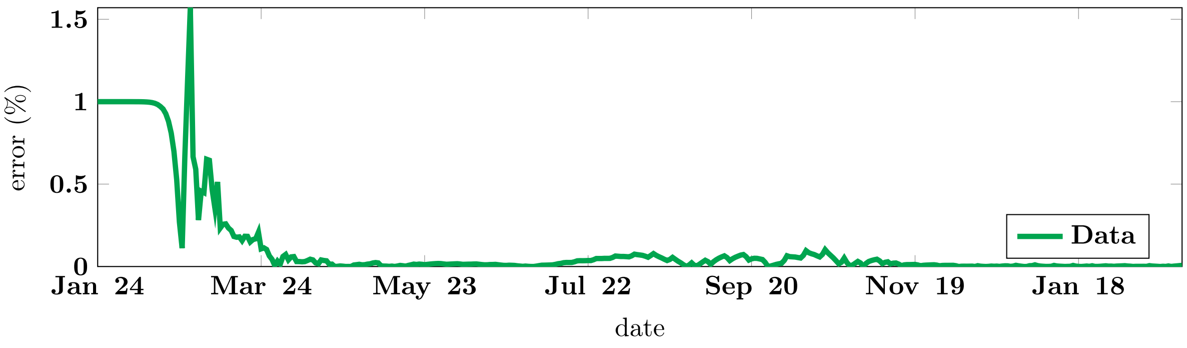


Figure 15: Relative error between the data and the model for France, expressed in percent.

#### 11.3 India

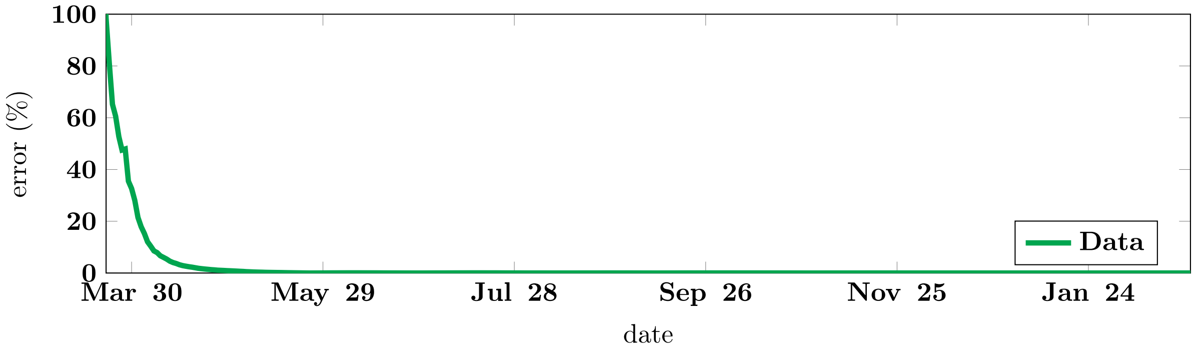


Figure 16: Relative error between the data and the model for India, expressed in percent.

#### 11.4 Israel

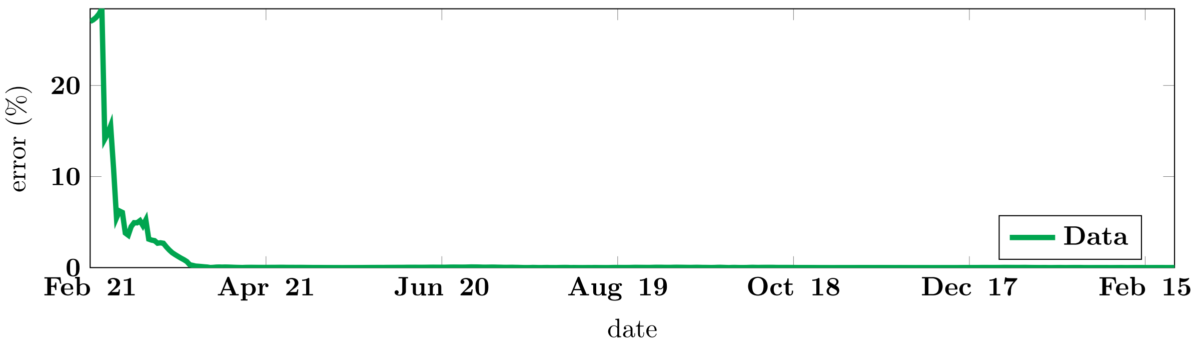


Figure 17: Relative error between the data and the model for Israel, expressed in percent.

#### 11.5 Japan

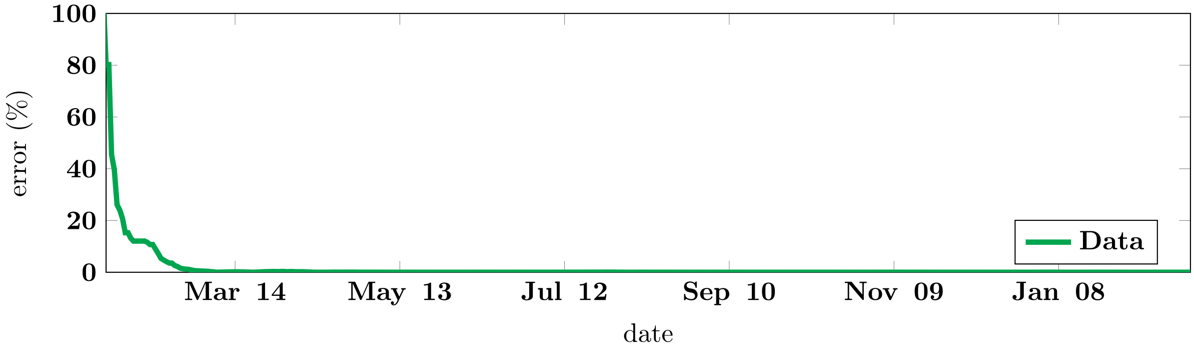


Figure 18: Relative error between the data and the model for Japan, expressed in percent.

#### 11.6 Peru

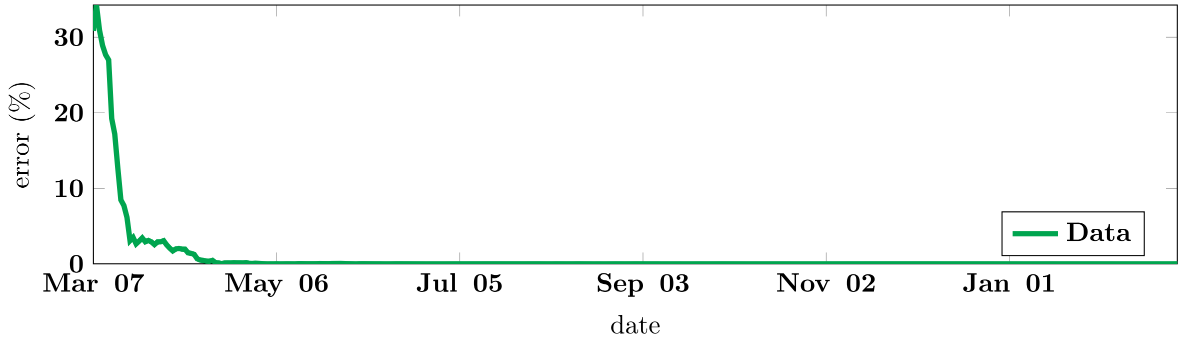


Figure 19: Relative error between the data and the model for Peru, expressed in percent.

#### 11.7 Spain

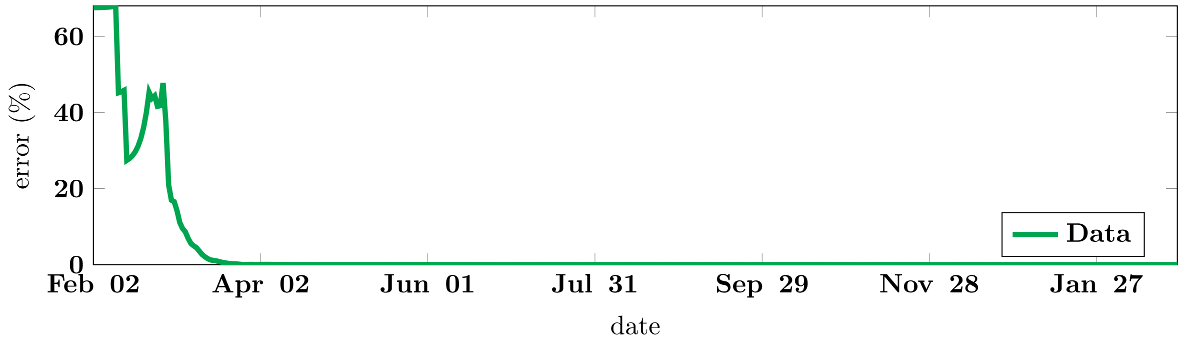


Figure 20: Relative error between the data and the model for Spain, expressed in percent.

## 11.8 UK

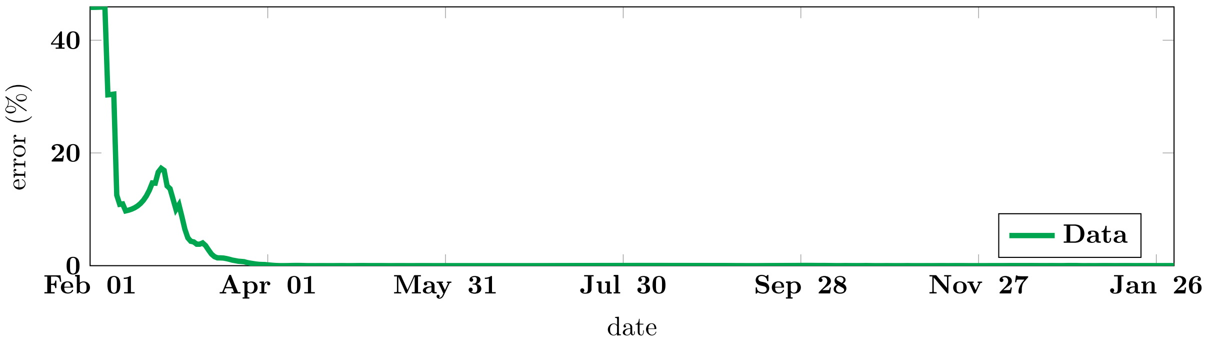


Figure 21: Relative error between the data and the model for UK, expressed in percent.

### 12 Table of estimated parameters for the phenomenological model

#### 12.1 California

| Period | Parameters value | Method | 95% Confidence interval |
|---|---|---|---|
| Period 1: Epidemic phase Mar 26, 2020 - Jun 11, 2020 | $N_0 = 7.34 \times 10^3$ | fitted | $N_0 \in [4.16 \times 10^3, 1.05 \times 10^4]$ |
| | $N_{base} = 1.14 \times 10^{-5}$ | fitted | $N_{base} \in [-4.33 \times 10^3, 4.33 \times 10^3]$ |
| | $N_\infty = 3.24 \times 10^5$ | fitted | $N_\infty \in [2.52 \times 10^5, 3.96 \times 10^5]$ |
| | $\chi = 4.14 \times 10^4$ | fitted | $\chi \in [7.74 \times 10^2, 8.20 \times 10^4]$ |
| | $\theta = 4.62 \times 10^{-7}$ | fitted | $\theta \in [2.39 \times 10^{-8}, 9.00 \times 10^{-7}]$ |
| Period 2: Endemic phase Jun 11, 2020 - Jun 23, 2020 | $a = 3.81 \times 10^3$ | computed | |
| | $N_0 = 1.36 \times 10^5$ | computed | |
| Period 3: Epidemic phase Jun 23, 2020 - Sep 20, 2020 | $N_0 = 1.57 \times 10^5$ | fitted | $N_0 \in [5.96 \times 10^4, 2.55 \times 10^5]$ |
| | $N_{base} = 2.45 \times 10^4$ | fitted | $N_{base} \in [-7.58 \times 10^4, 1.25 \times 10^5]$ |
| | $N_\infty = 8.22 \times 10^5$ | fitted | $N_\infty \in [7.36 \times 10^5, 9.08 \times 10^5]$ |
| | $\chi = 5.54 \times 10^{-2}$ | fitted | $\chi \in [5.32 \times 10^{-3}, 1.05 \times 10^{-1}]$ |
| | $\theta = 7.18 \times 10^{-1}$ | fitted | $\theta \in [-3.59 \times 10^{-2}, 1.47]$ |
| Period 4: Endemic phase Sep 20, 2020 - Nov 01, 2020 | $a = 3.66 \times 10^3$ | computed | |
| | $N_0 = 7.76 \times 10^5$ | computed | |
| Period 5: Epidemic phase Nov 01, 2020 - Feb 25, 2021 | $N_0 = 6.27 \times 10^4$ | fitted | $N_0 \in [4.95 \times 10^4, 7.59 \times 10^4]$ |
| | $N_{base} = 8.67 \times 10^5$ | fitted | $N_{base} \in [8.45 \times 10^5, 8.88 \times 10^5]$ |
| | $N_\infty = 2.66 \times 10^6$ | fitted | $N_\infty \in [2.64 \times 10^6, 2.67 \times 10^6]$ |
| | $\chi = 6.36 \times 10^{-2}$ | fitted | $\chi \in [5.73 \times 10^{-2}, 6.98 \times 10^{-2}]$ |
| | $\theta = 1.02$ | fitted | $\theta \in [8.79 \times 10^{-1}, 1.16]$ |

Table 10: *In this table we list the values of the parameters of the phenomenological model which give the best fit to the cumulative number of cases data in California from January 03 2020 to February 25 2021.*

#### 12.2 France

| Period | Parameters value | Method | 95% Confidence interval |
|---|---|---|---|
| Period 1: Epidemic phase Feb 27, 2020 - May 17, 2020 | $N_0 = 3.61 \times 10^{-4}$ | fitted | $N_0 \in [-3.77, 3.77]$ |
| | $N_{base} = 0.00$ | fixed | |
| | $N_\infty = 1.43 \times 10^5$ | fitted | $N_\infty \in [-1.58 \times 10^4, 3.01 \times 10^5]$ |
| | $\chi = 1.17 \times 10^2$ | fitted | $\chi \in [-1.09 \times 10^7, 1.09 \times 10^7]$ |
| | $\theta = 7.29 \times 10^{-4}$ | fitted | $\theta \in [-6.84 \times 10^1, 6.84 \times 10^1]$ |
| Period 2: Endemic phase May 17, 2020 - Jul 05, 2020 | $a = 3.14 \times 10^2$ | computed | |
| | $N_0 = 1.39 \times 10^5$ | computed | |
| Period 3: Epidemic phase Jul 05, 2020 - Nov 26, 2020 | $N_0 = 1.50 \times 10^4$ | fitted | $N_0 \in [1.36 \times 10^4, 1.65 \times 10^4]$ |
| | $N_{base} = 1.40 \times 10^5$ | fitted | $N_{base} \in [1.33 \times 10^5, 1.46 \times 10^5]$ |
| | $N_\infty = 1.99 \times 10^6$ | fitted | $N_\infty \in [1.97 \times 10^6, 2.01 \times 10^6]$ |
| | $\chi = 3.68 \times 10^{-2}$ | fitted | $\chi \in [3.60 \times 10^{-2}, 3.76 \times 10^{-2}]$ |
| | $\theta = 6.55$ | fitted | $\theta \in [5.52, 7.58]$ |
| Period 4: Endemic phase Nov 26, 2020 - Dec 20, 2020 | $a = 1.28 \times 10^4$ | computed | |
| | $N_0 = 2.11 \times 10^6$ | computed | |
| Period 5: Epidemic phase Dec 20, 2020 - Feb 25, 2021 | $N_0 = 2.73 \times 10^5$ | fitted | $N_0 \in [-2.43 \times 10^3, 5.48 \times 10^5]$ |
| | $N_{base} = 2.15 \times 10^6$ | fitted | $N_{base} \in [1.86 \times 10^6, 2.43 \times 10^6]$ |
| | $N_\infty = 2.13 \times 10^6$ | fitted | $N_\infty \in [1.88 \times 10^6, 2.39 \times 10^6]$ |
| | $\chi = 5.88 \times 10^{-2}$ | fitted | $\chi \in [-6.11 \times 10^{-2}, 1.79 \times 10^{-1}]$ |
| | $\theta = 5.47 \times 10^{-1}$ | fitted | $\theta \in [-9.19 \times 10^{-1}, 2.01]$ |

Table 11: *In this table we list the values of the parameters of the phenomenological model which give the best fit to the cumulative number of cases data in France from January 03 2020 to February 25 2021.*

#### 12.3 India

| Period | Parameters value | Method | 95% Confidence interval |
|---|---|---|---|
| Period 1: Epidemic phase<br>Feb 01, 2020 - Feb 25, 2021 | $N_0 = 5.83 \times 10^2$<br>$N_{base} = 1.97 \times 10^4$<br>$N_\infty = 1.10 \times 10^7$<br>$\chi = 4.89 \times 10^{-2}$<br>$\theta = 5.12 \times 10^{-1}$ | fitted<br>fitted<br>fitted<br>fitted<br>fitted | $N_0 \in [3.45 \times 10^2, 8.20 \times 10^2]$<br>$N_{base} \in [5.36 \times 10^3, 3.39 \times 10^4]$<br>$N_\infty \in [1.10 \times 10^7, 1.11 \times 10^7]$<br>$\chi \in [4.59 \times 10^{-2}, 5.20 \times 10^{-2}]$<br>$\theta \in [4.71 \times 10^{-1}, 5.54 \times 10^{-1}]$ |

Table 12: *In this table we list the values of the parameters of the phenomenological model which give the best fit to the cumulative number of cases data in India from January 03 2020 to February 25 2021.*

#### 12.4 Israel

| Period | Parameters value | Method | 95% Confidence interval |
|---|---|---|---|
| Period 1: Epidemic phase<br>Feb 27, 2020 - Jun 01, 2020 | $N_0 = 1.08 \times 10^{-2}$<br>$N_{base} = 4.27 \times 10^1$<br>$N_\infty = 1.71 \times 10^4$<br>$\chi = 9.18 \times 10^{-1}$<br>$\theta = 1.05 \times 10^{-1}$ | fitted<br>fitted<br>fitted<br>fitted<br>fitted | $N_0 \in [-3.85 \times 10^{-2}, 6.02 \times 10^{-2}]$<br>$N_{base} \in [-3.36 \times 10^1, 1.19 \times 10^2]$<br>$N_\infty \in [1.70 \times 10^4, 1.72 \times 10^4]$<br>$\chi \in [1.71 \times 10^{-1}, 1.67]$<br>$\theta \in [1.55 \times 10^{-2}, 1.94 \times 10^{-1}]$ |
| Period 2: Endemic phase<br>Jun 01, 2020 - Jun 25, 2020 | $a = 2.04 \times 10^2$<br>$N_0 = 1.70 \times 10^4$ | computed<br>computed | |
| Period 3: Epidemic phase<br>Jun 25, 2020 - Aug 08, 2020 | $N_0 = 2.48 \times 10^3$<br>$N_{base} = 1.95 \times 10^4$<br>$N_\infty = 8.66 \times 10^4$<br>$\chi = 2.93 \times 10^{-1}$<br>$\theta = 2.04 \times 10^{-1}$ | fitted<br>fitted<br>fitted<br>fitted<br>fitted | $N_0 \in [3.43 \times 10^2, 4.61 \times 10^3]$<br>$N_{base} \in [1.70 \times 10^4, 2.20 \times 10^4]$<br>$N_\infty \in [7.78 \times 10^4, 9.55 \times 10^4]$<br>$\chi \in [-2.61 \times 10^{-1}, 8.48 \times 10^{-1}]$<br>$\theta \in [-2.43 \times 10^{-1}, 6.50 \times 10^{-1}]$ |
| Period 4: Endemic phase<br>Aug 08, 2020 - Sep 03, 2020 | $a = 1.54 \times 10^3$<br>$N_0 = 7.97 \times 10^4$ | computed<br>computed | |
| Period 5: Epidemic phase<br>Sep 03, 2020 - Oct 20, 2020 | $N_0 = 4.59 \times 10^4$<br>$N_{base} = 7.38 \times 10^4$<br>$N_\infty = 2.35 \times 10^5$<br>$\chi = 5.05 \times 10^{-2}$<br>$\theta = 3.45$ | fitted<br>fitted<br>fitted<br>fitted<br>fitted | $N_0 \in [2.88 \times 10^4, 6.31 \times 10^4]$<br>$N_{base} \in [5.53 \times 10^4, 9.23 \times 10^4]$<br>$N_\infty \in [2.19 \times 10^5, 2.52 \times 10^5]$<br>$\chi \in [3.77 \times 10^{-2}, 6.34 \times 10^{-2}]$<br>$\theta \in [1.96, 4.93]$ |
| Period 6: Endemic phase<br>Oct 20, 2020 - Nov 14, 2020 | $a = 8.90 \times 10^2$<br>$N_0 = 3.04 \times 10^5$ | computed<br>computed | |
| Period 7: Epidemic phase<br>Nov 14, 2020 - Feb 25, 2021 | $N_0 = 3.16 \times 10^3$<br>$N_{base} = 3.23 \times 10^5$<br>$N_\infty = 4.87 \times 10^5$<br>$\chi = 8.28 \times 10^{-2}$<br>$\theta = 7.06 \times 10^{-1}$ | fitted<br>fitted<br>fitted<br>fitted<br>fitted | $N_0 \in [2.16 \times 10^3, 4.17 \times 10^3]$<br>$N_{base} \in [3.21 \times 10^5, 3.25 \times 10^5]$<br>$N_\infty \in [4.79 \times 10^5, 4.95 \times 10^5]$<br>$\chi \in [7.22 \times 10^{-2}, 9.34 \times 10^{-2}]$<br>$\theta \in [5.69 \times 10^{-1}, 8.43 \times 10^{-1}]$ |

Table 13: *In this table we list the values of the parameters of the phenomenological model which give the best fit to the cumulative number of cases data in Israel from January 03 2020 to February 25 2021.*

#### 12.5 Japan

| Period | Parameters value | Method | 95% Confidence interval |
|---|---|---|---|
| Period 1: Epidemic phase Feb 20, 2020 - May 27, 2020 | $N_0 = 5.83$ | fitted | $N_0 \in [1.91, 9.74]$ |
| | $N_{base} = 3.25 \times 10^2$ | fitted | $N_{base} \in [2.55 \times 10^2, 3.95 \times 10^2]$ |
| | $N_\infty = 1.63 \times 10^4$ | fitted | $N_\infty \in [1.62 \times 10^4, 1.64 \times 10^4]$ |
| | $\chi = 1.48 \times 10^{-1}$ | fitted | $\chi \in [1.30 \times 10^{-1}, 1.65 \times 10^{-1}]$ |
| | $\theta = 8.29 \times 10^{-1}$ | fitted | $\theta \in [6.88 \times 10^{-1}, 9.70 \times 10^{-1}]$ |
| Period 2: Endemic phase May 27, 2020 - Jun 13, 2020 | $a = 7.07 \times 10^1$ | computed | |
| | $N_0 = 1.65 \times 10^4$ | computed | |
| Period 3: Epidemic phase Jun 13, 2020 - Sep 10, 2020 | $N_0 = 1.49 \times 10^2$ | fitted | $N_0 \in [8.52 \times 10^1, 2.13 \times 10^2]$ |
| | $N_{base} = 1.75 \times 10^4$ | fitted | $N_{base} \in [1.73 \times 10^4, 1.78 \times 10^4]$ |
| | $N_\infty = 6.02 \times 10^4$ | fitted | $N_\infty \in [5.93 \times 10^4, 6.10 \times 10^4]$ |
| | $\chi = 1.19 \times 10^{-1}$ | fitted | $\chi \in [1.03 \times 10^{-1}, 1.35 \times 10^{-1}]$ |
| | $\theta = 6.28 \times 10^{-1}$ | fitted | $\theta \in [5.04 \times 10^{-1}, 7.52 \times 10^{-1}]$ |
| Period 4: Endemic phase Sep 10, 2020 - Oct 18, 2020 | $a = 5.36 \times 10^2$ | computed | |
| | $N_0 = 7.27 \times 10^4$ | computed | |
| Period 5: Epidemic phase Oct 18, 2020 - Dec 05, 2020 | $N_0 = 6.33 \times 10^3$ | fitted | $N_0 \in [4.64 \times 10^3, 8.01 \times 10^3]$ |
| | $N_{base} = 8.68 \times 10^4$ | fitted | $N_{base} \in [8.48 \times 10^4, 8.88 \times 10^4]$ |
| | $N_\infty = 9.10 \times 10^4$ | fitted | $N_\infty \in [7.75 \times 10^4, 1.05 \times 10^5]$ |
| | $\chi = 5.60 \times 10^{-2}$ | fitted | $\chi \in [4.74 \times 10^{-2}, 6.46 \times 10^{-2}]$ |
| | $\theta = 2.58$ | fitted | $\theta \in [1.00, 4.16]$ |
| Period 6: Epidemic phase Dec 05, 2020 - Dec 30, 2020 | $N_0 = 1.23 \times 10^5$ | fitted | $N_0 \in [-2.43 \times 10^5, 4.90 \times 10^5]$ |
| | $N_{base} = 3.43 \times 10^4$ | fitted | $N_{base} \in [-3.33 \times 10^5, 4.01 \times 10^5]$ |
| | $N_\infty = 3.49 \times 10^5$ | fitted | $N_\infty \in [-2.92 \times 10^7, 2.99 \times 10^7]$ |
| | $\chi = 1.78 \times 10^{-2}$ | fitted | $\chi \in [-3.59 \times 10^{-2}, 7.15 \times 10^{-2}]$ |
| | $\theta = 7.84$ | fitted | $\theta \in [-1.28 \times 10^3, 1.30 \times 10^3]$ |
| Period 7: Epidemic phase Dec 30, 2020 - Feb 25, 2021 | $N_0 = 2.00 \times 10^4$ | fitted | $N_0 \in [1.59 \times 10^3, 3.84 \times 10^4]$ |
| | $N_{base} = 2.05 \times 10^5$ | fitted | $N_{base} \in [1.85 \times 10^5, 2.25 \times 10^5]$ |
| | $N_\infty = 2.29 \times 10^5$ | fitted | $N_\infty \in [2.11 \times 10^5, 2.47 \times 10^5]$ |
| | $\chi = 7.98 \times 10^{-1}$ | fitted | $\chi \in [-2.54, 4.13]$ |
| | $\theta = 9.61 \times 10^{-2}$ | fitted | $\theta \in [-3.15 \times 10^{-1}, 5.07 \times 10^{-1}]$ |

Table 14: *In this table we list the values of the parameters of the phenomenological model which give the best fit to the cumulative number of cases data in Japan from January 03 2020 to February 25 2021.*

#### 12.6 Peru

| Period | Parameters value | Method | 95% Confidence interval |
|---|---|---|---|
| Period 1: Epidemic phase Mar 20, 2020 - Jul 01, 2020 | $N_0 = 8.36 \times 10^2$ | fitted | $N_0 \in [2.63 \times 10^2, 1.41 \times 10^3]$ |
| | $N_{base} = 3.00 \times 10^{-5}$ | fitted | $N_{base} \in [-1.74 \times 10^3, 1.74 \times 10^3]$ |
| | $N_\infty = 3.61 \times 10^5$ | fitted | $N_\infty \in [3.44 \times 10^5, 3.79 \times 10^5]$ |
| | $\chi = 1.08 \times 10^{-1}$ | fitted | $\chi \in [7.59 \times 10^{-2}, 1.41 \times 10^{-1}]$ |
| | $\theta = 4.20 \times 10^{-1}$ | fitted | $\theta \in [2.41 \times 10^{-1}, 5.98 \times 10^{-1}]$ |
| Period 2: Endemic phase Jul 01, 2020 - Jul 30, 2020 | $a = 3.67 \times 10^3$ | computed | |
| | $N_0 = 2.83 \times 10^5$ | computed | |
| Period 3: Epidemic phase Jul 30, 2020 - Nov 10, 2020 | $N_0 = 1.86 \times 10^5$ | fitted | $N_0 \in [-2.61 \times 10^4, 3.98 \times 10^5]$ |
| | $N_{base} = 2.03 \times 10^5$ | fitted | $N_{base} \in [-1.11 \times 10^4, 4.18 \times 10^5]$ |
| | $N_\infty = 7.69 \times 10^5$ | fitted | $N_\infty \in [5.65 \times 10^5, 9.72 \times 10^5]$ |
| | $\chi = 4.84 \times 10^{-1}$ | fitted | $\chi \in [-6.23, 7.20]$ |
| | $\theta = 5.95 \times 10^{-2}$ | fitted | $\theta \in [-7.74 \times 10^{-1}, 8.93 \times 10^{-1}]$ |
| Period 4: Endemic phase Nov 10, 2020 - Jan 11, 2021 | $a = 1.80 \times 10^3$ | computed | |
| | $N_0 = 9.16 \times 10^5$ | computed | |
| Period 5: Epidemic phase Jan 11, 2021 - Feb 25, 2021 | $N_0 = 3.23 \times 10^5$ | fitted | |
| | $N_{base} = 7.04 \times 10^5$ | fitted | |
| | $N_\infty = 7.00 \times 10^6$ | fitted | |
| | $\chi = 1.36 \times 10^{-2}$ | fitted | |
| | $\theta = 3.67 \times 10^1$ | fitted | |

Table 15: *In this table we list the values of the parameters of the phenomenological model which give the best fit to the cumulative number of cases data in Peru from January 03 2020 to February 25 2021.*

#### 12.7 Spain

| Period | Parameters value | Method | 95% Confidence interval |
|---|---|---|---|
| Period 1: Epidemic phase<br>Feb 15, 2020 - May 10, 2020 | $N_0 = 5.19 \times 10^{-4}$<br>$N_{base} = 5.77 \times 10^2$<br>$N_\infty = 2.32 \times 10^5$<br>$\chi = 9.80 \times 10^{-1}$<br>$\theta = 9.75 \times 10^{-2}$ | fitted<br>fitted<br>fitted<br>fitted<br>fitted | $N_0 \in [-5.00 \times 10^{-3}, 6.04 \times 10^{-3}]$<br>$N_{base} \in [-4.50 \times 10^2, 1.60 \times 10^3]$<br>$N_\infty \in [2.30 \times 10^5, 2.34 \times 10^5]$<br>$\chi \in [-1.26 \times 10^{-1}, 2.09]$<br>$\theta \in [-1.83 \times 10^{-2}, 2.13 \times 10^{-1}]$ |
| Period 2: Endemic phase<br>May 10, 2020 - Jun 22, 2020 | $a = 5.67 \times 10^2$<br>$N_0 = 2.28 \times 10^5$ | computed<br>computed | |
| Period 3: Epidemic phase<br>Jun 22, 2020 - Oct 02, 2020 | $N_0 = 2.38 \times 10^3$<br>$N_{base} = 2.50 \times 10^5$<br>$N_\infty = 9.89 \times 10^5$<br>$\chi = 9.29 \times 10^{-2}$<br>$\theta = 3.84 \times 10^{-1}$ | fitted<br>fitted<br>fitted<br>fitted<br>fitted | $N_0 \in [1.39 \times 10^3, 3.36 \times 10^3]$<br>$N_{base} \in [2.48 \times 10^5, 2.53 \times 10^5]$<br>$N_\infty \in [9.02 \times 10^5, 1.08 \times 10^6]$<br>$\chi \in [7.07 \times 10^{-2}, 1.15 \times 10^{-1}]$<br>$\theta \in [2.38 \times 10^{-1}, 5.29 \times 10^{-1}]$ |
| Period 4: Endemic phase<br>Oct 02, 2020 - Oct 18, 2020 | $a = 1.09 \times 10^4$<br>$N_0 = 8.14 \times 10^5$ | computed<br>computed | |
| Period 5: Epidemic phase<br>Oct 18, 2020 - Dec 06, 2020 | $N_0 = 1.68 \times 10^5$<br>$N_{base} = 8.20 \times 10^5$<br>$N_\infty = 9.85 \times 10^5$<br>$\chi = 3.15 \times 10^{-1}$<br>$\theta = 2.02 \times 10^{-1}$ | fitted<br>fitted<br>fitted<br>fitted<br>fitted | $N_0 \in [-3.50 \times 10^4, 3.72 \times 10^5]$<br>$N_{base} \in [6.12 \times 10^5, 1.03 \times 10^6]$<br>$N_\infty \in [8.01 \times 10^5, 1.17 \times 10^6]$<br>$\chi \in [-1.05, 1.68]$<br>$\theta \in [-7.15 \times 10^{-1}, 1.12]$ |
| Period 6: Endemic phase<br>Dec 06, 2020 - Dec 26, 2020 | $a = 9.15 \times 10^3$<br>$N_0 = 1.72 \times 10^6$ | computed<br>computed | |
| Period 7: Epidemic phase<br>Dec 26, 2020 - Feb 25, 2021 | $N_0 = 5.94 \times 10^4$<br>$N_{base} = 1.84 \times 10^6$<br>$N_\infty = 1.30 \times 10^6$<br>$\chi = 1.30 \times 10^{-1}$<br>$\theta = 7.84 \times 10^{-1}$ | fitted<br>fitted<br>fitted<br>fitted<br>fitted | $N_0 \in [3.86 \times 10^4, 8.02 \times 10^4]$<br>$N_{base} \in [1.81 \times 10^6, 1.87 \times 10^6]$<br>$N_\infty \in [1.28 \times 10^6, 1.32 \times 10^6]$<br>$\chi \in [9.90 \times 10^{-2}, 1.60 \times 10^{-1}]$<br>$\theta \in [5.50 \times 10^{-1}, 1.02]$ |

Table 16: *In this table we list the values of the parameters of the phenomenological model which give the best fit to the cumulative number of cases data in Spain from January 03 2020 to February 01 2021.*

#### 12.8 United Kingdom

| Period | Parameters value | Method | 95% Confidence interval |
|---|---|---|---|
| Period 1: Epidemic phase<br>Feb 15, 2020 - Jun 15, 2020 | $N_0 = 2.65 \times 10^{-2}$<br>$N_{base} = 1.12 \times 10^2$<br>$N_\infty = 2.86 \times 10^5$<br>$\chi = 1.76$<br>$\theta = 2.76 \times 10^{-2}$ | fitted<br>fitted<br>fitted<br>fitted<br>fitted | $N_0 \in [-8.82 \times 10^{-2}, 1.41 \times 10^{-1}]$<br>$N_{base} \in [-4.82 \times 10^2, 7.06 \times 10^2]$<br>$N_\infty \in [2.84 \times 10^5, 2.88 \times 10^5]$<br>$\chi \in [-1.46, 4.98]$<br>$\theta \in [-2.38 \times 10^{-2}, 7.90 \times 10^{-2}]$ |
| Period 2: Endemic phase<br>Jun 15, 2020 - Sep 01, 2020 | $a = 9.43 \times 10^2$<br>$N_0 = 2.70 \times 10^5$ | computed<br>computed | |
| Period 3: Epidemic phase<br>Sep 01, 2020 - Nov 20, 2020 | $N_0 = 7.85 \times 10^3$<br>$N_{base} = 3.36 \times 10^5$<br>$N_\infty = 2.14 \times 10^6$<br>$\chi = 2.41 \times 10^{-1}$<br>$\theta = 1.32 \times 10^{-1}$ | fitted<br>fitted<br>fitted<br>fitted<br>fitted | $N_0 \in [3.63 \times 10^3, 1.21 \times 10^4]$<br>$N_{base} \in [3.28 \times 10^5, 3.43 \times 10^5]$<br>$N_\infty \in [1.93 \times 10^6, 2.36 \times 10^6]$<br>$\chi \in [2.16 \times 10^{-2}, 4.60 \times 10^{-1}]$<br>$\theta \in [-9.25 \times 10^{-3}, 2.74 \times 10^{-1}]$ |
| Period 4: Endemic phase<br>Nov 20, 2020 - Dec 10, 2020 | $a = 1.61 \times 10^4$<br>$N_0 = 1.48 \times 10^6$ | computed<br>computed | |
| Period 5: Epidemic phase<br>Dec 10, 2020 - Feb 01, 2021 | $N_0 = 2.26 \times 10^5$<br>$N_{base} = 1.58 \times 10^6$<br>$N_\infty = 2.42 \times 10^6$<br>$\chi = 8.57 \times 10^{-2}$<br>$\theta = 1.08$ | fitted<br>fitted<br>fitted<br>fitted<br>fitted | $N_0 \in [1.16 \times 10^5, 3.35 \times 10^5]$<br>$N_{base} \in [1.46 \times 10^6, 1.70 \times 10^6]$<br>$N_\infty \in [2.34 \times 10^6, 2.51 \times 10^6]$<br>$\chi \in [5.14 \times 10^{-2}, 1.20 \times 10^{-1}]$<br>$\theta \in [4.85 \times 10^{-1}, 1.68]$ |

Table 17: *In this table we list the values of the parameters of the phenomenological model which give the best fit to the cumulative number of cases data in United Kingdom from January 03 2020 to February 01 2021.*

### 13 Stochastic approach to effective reproductive ratio

In numerical applications, we also present the results obtained by applying the method described in the paper of Cori et al. [8]. Let us summarize the principle of the method. We consider that the incidence data (*i.e.* the daily number of new reported cases) correspond to infection events that have occurred in the past. For each new reported case we reconstruct the time at which the infectious period started by sampling a Gamma distribution (i.e. the time from the infection to the moment at which the individual are reported follows a Gamma distribution). The parameters of this Gamma distribution are computed to match the differential equation framework. In numerical application, $1/\mu = 10$ days we took the average for the average of the Gamma distribution as well as its standard deviation. We denote $I_t$ the resulting number of individuals that begin their infectious period on day $t$. As described in [8], we use a smoothing window of $\tau$ days ($\tau = 14$ days in numerical applications). The resulting effective reproductive ratio $R_t$ is then computed as

$$R_t = \frac{a + \displaystyle\sum_{s=t-\tau+1}^{t} I_s}{\frac{1}{b} + \sum_{s=t-\tau+1}^{t} \Lambda_s},$$

where $a$ and $b$ are a prior distribution on $R_t$ (we took $a = 1$ and $b = 5$, as in [8]) and $\Lambda_s$ is computed by the formula

$$\Lambda_s = \sum_{s=1}^{t} I_{t-s} w_s,$$

where $w_s$ is the average infectiousness profile after time $s$. In numerical applications, and following [8, Web Appendix 11], we used the following formula for $w_s$

$$\begin{aligned} w_s &= sF_{\Gamma,\alpha,\beta}(s) + (s-2)F_{\Gamma,\alpha,\beta}(s-2) - 2(s-1)F_{\Gamma,\alpha,\beta}(s-1) \\ &\quad + \alpha\beta(2F_{\Gamma,\alpha+1,\beta}(s-1) - F_{\Gamma,\alpha+1,\beta}(s-2) - F_{\Gamma,\alpha+1,\beta}(s)), \end{aligned}$$

where $F_{\Gamma,\alpha,\beta}(s)$ is the cumulative density of a Gamma distribution of parameters $(\alpha, \beta)$:

$$F_{\Gamma,\alpha,\beta}(t) = \int_0^t \frac{1}{\Gamma(\alpha)\beta^\alpha} s^{\alpha-1} e^{-\frac{s}{\beta}} \mathrm{d}s.$$

The parameters $\alpha$ and $\beta$ are computed to match the Gamma distribution of the serial intervals which, in our case, have mean value $1/\mu = 10$ days and standard deviation as well of $1/\mu$, so that $\alpha = 1/\mu$ and $\beta = 1/\mu$.

Because of the sampling of random numbers involved in the computation of $R_t$, the procedure described above was repeated 100 times (each time drawing a new sequence of $I_s$ from the daily number of new cases) and the final value of $R_t$ presented in Figure 4 of the main text (green curves) is the average of the values obtained during these 100 simulations.

### 14 Upper bound of the duration for the exposed period and the asymptomatic infectious period

Let us finally mention that for each country and each epidemic wave we evaluated the parameter $1/(\chi\,\theta)$. In Figure 22 we plot the histogram of its estimated value and obtain a median value be 15.61 days. Therefore the length of exposure and the length asymptomatic infectious period should smaller than 15.61 days.

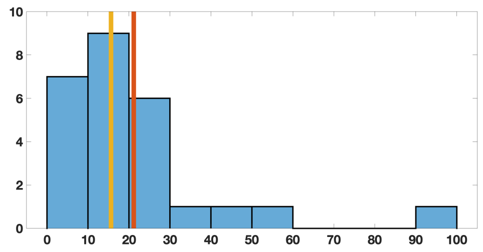


Figure 22: *In this Figure we plot the histogram for the estimated values* $1/(\chi\,\theta)$ *(see Appendix E). The red vertical line is mean value which is equal to* 21 *days. The yellow vertical line is median value which is equal to* 15.61 *days.*

In this section, we plot the estimated values of the parameter $1/(\chi\,\theta)$ for each epidemic period and each country consider in this study. The parameter corresponds to the upper bound of the length of the exposed period and asymptomatic infectious period. Indeed from the section devoted to the compatibility condition we know that the average duration of the exposed period should satisfy

$$1/\nu \leq 1/(\chi\,\theta),$$

and the average duration of the asymptomatic infectious period should should satisfy

$$1/\alpha \leq 1/(\chi\,\theta).$$

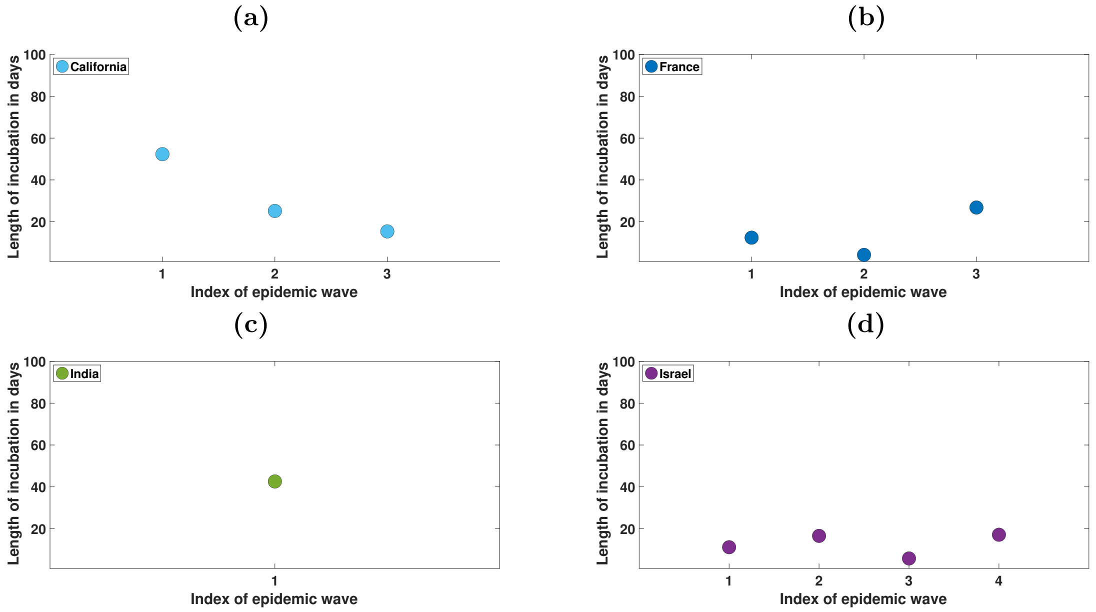


Figure 23: *In this figure we plot the values of the parameter* $1/(\chi\,\theta)$ *estimated for each epidemic wave and for California (a), France (b), India (c), Israel (d). This parameter represents the maximal length of the incubation period. In each figure, we plot this parameter for each epidemic wave and for each country.*

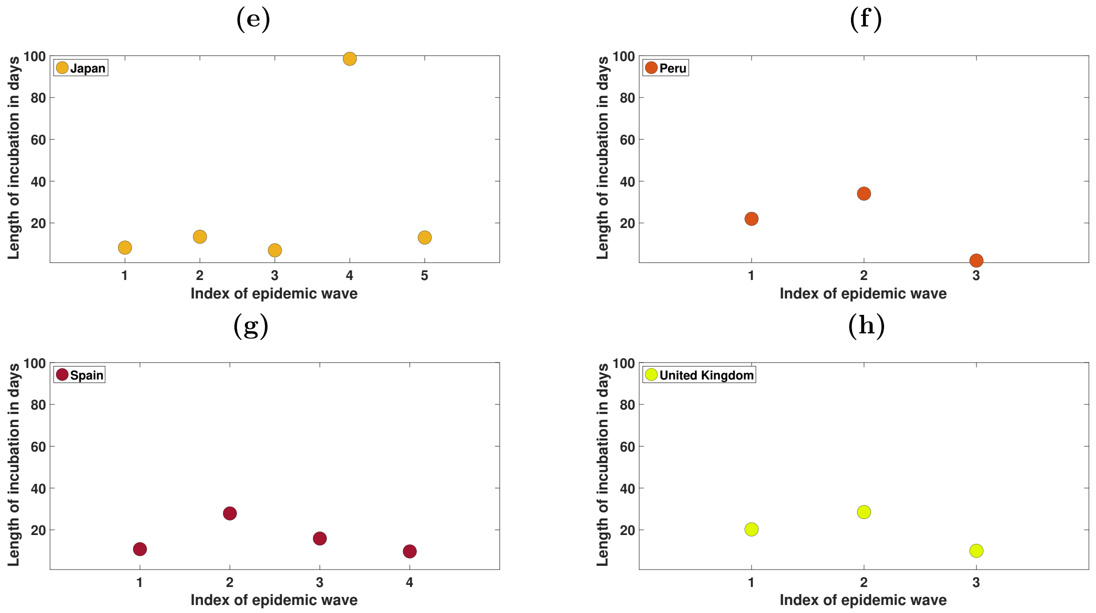


Figure 24: *In this figure we plot the values of the parameter* $1/(\chi\,\theta)$ *estimated for each epidemic wave and for Japan (e), Peru (f), Spain (g) and United Kingdom (h). This parameter represents the maximal length of the incubation period. In each figure, we plot this parameter for each epidemic wave and for each country.*

[3] D. Bernoulli. Essai d'une nouvelle analyse de la petite Vérole, & des avantages de l'Inoculation pour la prévenir. *Histoire de l'Académie royale des sciences avec les mémoires de mathématique et de physique tirés de cette Académie* **Année 1760.Mémoires** (1766). Disponible sur Gallica, pp. 1–45.

[9] O. Diekmann, H. Heesterbeek, and T. Britton. *Mathematical tools for understanding infectious disease dynamics*. Princeton Series in Theoretical and Computational Biology. Princeton University Press, Princeton, NJ, 2013, pp. xiv+502.

[16] M. J. Keeling and P. Rohani. *Modeling infectious diseases in humans and animals.* Princeton University Press, Princeton, NJ, 2008, pp. xvi+368.

[17] J. D. Murray. *Mathematical biology.* Second. Vol. 19. Biomathematics. Springer-Verlag, Berlin, 1993, pp. xiv+767.

[18] J. Scire et al. Reproductive number of the COVID-19 epidemic in Switzerland with a focus on the Cantons of Basel-Stadt and Basel-Landschaft. *Swiss Med. Wkly.* **150.19-20** (2020), w20271. DOI: 10.4414/smw.2020.20271.

[19] B. Tang et al. Estimation of the transmission risk of the 2019-nCoV and its implication for public health interventions. *J. Clin. Med.* **9.2** (2020), p. 462. DOI: 10.3390/jcm9020462.

[20] H. R. Thieme. *Mathematics in population biology.* Princeton Series in Theoretical and Computational Biology. Princeton University Press, Princeton, NJ, 2003, pp. xx+543.

[21] P. F. Verhulst. Notice sur la loi que la population poursuit dans son accroissement. *Correspondance mathématique et physique* **10** (1838), pp. 113–121.

[22] X.-S. Wang, J. Wu, and Y. Yang. Richards model revisited: validation by and application to infection dynamics. *J. Theoret. Biol.* **313** (2012), pp. 12–19. DOI: 10.1016/j.jtbi.2012.07.024.

[23] J. T. Wu, K. Leung, and G. M. Leung. Nowcasting and forecasting the potential domestic and international spread of the 2019-nCoV outbreak originating in Wuhan, China: a modelling study. *Lancet* **395.10225** (2020), pp. 689–697. DOI: 10.1016/S0140-6736(20)30260-9.

[24] G. Zhou and G. Yan. Severe acute respiratory syndrome epidemic in Asia. *Emerg. Infect. Dis.* **9.12** (2003), pp. 1608–1610.
